# Psychiatric symptom improvement from adjunctive statin prescribing in severe mental illness: three target trial emulation studies

**DOI:** 10.1101/2025.01.20.25320829

**Authors:** Naomi Launders, Alvin Richards-Belle, Kenneth KC Man, Ian CK Wong, David PJ Osborn, Joseph F Hayes

## Abstract

**Background:** Randomized controlled trials (RCTs) of statins as adjunct therapy for severe mental illness (SMI) have produced mixed results. Specific statin-antipsychotic combinations might improve psychiatric symptoms through: 1) blood-brain barrier (BBB) penetrant statins being anti-inflammatory/neuroprotective, and/or 2) statins that inhibit p-glycoprotein enhancing the effects of antipsychotics with high p-glycoprotein affinity.

**Aim:** To investigate these mechanisms via three target trial emulation studies.

**Methods:** We identified patients with SMI (schizophrenia, bipolar disorder, ‘other’ psychoses) prescribed antipsychotics/mood stabilisers and statins from 2000-2019 in English electronic health records. We defined three hypothetical RCTs and their observational analogues: 1) simvastatin (crosses BBB) vs. atorvastatin/pravastatin/rosuvastatin (non-penetrant); 2A) simvastatin/atorvastatin (p-glycoprotein inhibitors) vs. pravastatin in patients prescribed risperidone/olanzapine/aripiprazole (high p-glycoprotein affinity); 2B) risperidone/olanzapine/aripiprazole vs. quetiapine (low p-glycoprotein affinity) in patients prescribed simvastatin/atorvastatin. Primary outcome: 12-month psychiatric admissions. Secondary outcomes: self-harm, physical health, and accident/injury admissions.

**Results:** In 72,096 patients prescribed statins and antipsychotics/mood stabilisers, we found no reduction in psychiatric admissions at 12 months in patients prescribed: 1) BBB-penetrant vs. non-penetrant statins (HR:1.07; 95%CI:0.88-1.31); 2A) antipsychotics with p-glycoprotein affinity and p-glycoprotein inhibiting statins vs. statins without inhibition (HR:0.77; 95%CI:0.28-2.15); 2B) p-glycoprotein inhibiting statins with antipsychotics having p-glycoprotein affinity vs. antipsychotics without affinity (HR:0.93; 95%CI:0.79-1.09). In 2B we observed reduced self-harm (HR:0.60; 95%CI:0.38-0.97) in per-protocol analysis and reduced psychiatric admissions in the ‘other’ psychoses subgroup (HR:0.53; 95%CI:0.34-0.85).

**Conclusions:** BBB permeability is unlikely to be the mechanism by which statins improve SMI symptoms. Further research is needed to understand statin-antipsychotic interactions, and whether interaction with p-glycoprotein is a plausible mechanism.

## Introduction

In the UK, five statins (atorvastatin, fluvastatin, pravastatin, rosuvastatin and simvastatin), or hydroxylmethyl glutaryl coenzyme A reductase inhibitors, are routinely prescribed to reduce cholesterol levels (Grundy et al., 2019). It has been hypothesised that, given the anti-inflammatory effect of statins and the potential role of inflammation in mental illness, statins may improve psychiatric symptoms (Shen et al., 2018; Sommer et al., 2021; Postolache et al., 2021) and may have neuroprotective effects (Sierra et al., 2011).

Statins have been investigated as adjunct therapy for a range of neuropsychiatric disorders, including depression (Xiao et al., 2023) and severe mental illness (SMI; schizophrenia, bipolar disorder and non-organic non-affective psychoses) (Avan et al., 2021). However, the results of both randomised controlled trials (RCTs) and observational studies have been mixed. Two meta-analyses of RCTs reported a modest improvement in the Positive and Negative Syndrome Scale (PANSS) total scores in people with schizophrenia prescribed statins as an adjunct (Nomura et al., 2018; Shen et al., 2018).

However, concern has been raised about the heterogeneity of the originating studies included in these meta-analyses in terms of stage of illness, the combination of both end-point and change scores, and the size and generalisability of the RCT populations included (Andrade, 2018). A more recent meta-analysis found that statins improved total PANSS scores and negative sub-scales in people with schizophrenia, but not positive PANSS scales (Peng et al., 2024).

Individual RCTs have found varying results and have included small numbers of patients (Sommer et al., 2021; Tajik-Esmaeeli et al., 2017; Chaudhry et al., 2014; Aichholzer et al., 2022; Vincenzi et al., 2014; Ghanizadeh et al., 2014; Sayyah et al., 2015; Weiser et al., 2023). Several observational studies have found lower risk of psychiatric admissions in people diagnosed with SMI prescribed statins. A study from the US found a lower risk of psychiatric admissions in those prescribed statins (schizophrenia relative risk: 0.81 for lipophilic statins; 95%CI:0.77-0.86 and 0.73; 95%CI:0.67-0.80 with hydrophilic statins), though noted that confounding by indication could account for these findings (Postolache et al., 2021). A self-controlled study by Hayes et al. found that patients with SMI were less likely to experience psychiatric admissions during periods of statin prescriptions overall (HR: 0.86, 95%CI: 0.83-0.89), and when stratified by SMI diagnosis (Hayes et al., 2019). Recently, a study of patients with schizophrenia in Taiwan found that while lipid-lowering agents were associated with lower all-cause mortality, there was no association with suicide (Chen et al., 2024).

Given the heterogenous results regarding the potential for statins to reduce psychiatric symptoms, and the proposed mechanisms for this, it seems likely that any improvements may be dependent either on patient characteristics, or the combination of statin and antipsychotic or mood stabilisers prescribed. Despite this, no studies have investigated the comparative effect of common combinations of antipsychotics or mood stabilisers and statins.

If the primary mechanism is via dampening central nervous system inflammation, statins which cross the blood brain barrier (BBB) more readily (i.e., lipophilic statins) would theoretically have a greater anti-inflammatory, antioxidant and neuroprotective effect (Postolache et al., 2021; Sierra et al., 2011). However, Postolache et al. compared hydrophilic statins (rosuvastatin, pravastatin, and fluvastatin) to lipophilic statins (simvastatin, atorvastatin, pitavastatin, and lovastatin) in patients with bipolar disorder or schizophrenia and found those prescribed hydrophilic statins had fewer admissions (Postolache et al., 2021). Importantly, lipophilicity is not the only predictor of BBB penetrance: the larger molecular weight of atorvastatin and rosuvastatin may hinder BBB transfer (Sierra et al., 2011). A study by Sierra et al. found that atorvastatin, rosuvastatin and pravastatin do not readily cross the BBB by passive diffusion, while simvastatin crossed most readily, followed by fluvastatin (Sierra et al., 2011).

A second hypothesised mechanism of action is through improved effectiveness of antipsychotics. The membrane protein P-glycoprotein may be involved in the cellular uptake and efflux of some antipsychotics. Aripiprazole, risperidone and olanzapine have affinity for the P-glycoprotein in vitro, with higher brain concentrations of these antipsychotics in animal models where P-glycoprotein is absent or inhibited. Risperidone concentration in humans is higher in the presence of P-glycoprotein inhibitors (Moons et al., 2011). Simvastatin and atorvastatin may act as inhibitors of P-glycoprotein, thereby potentially modulating the concentration of antipsychotics such as aripiprazole, risperidone and olanzapine, but not quetiapine (Holtzman et al., 2006).

We aimed to complete three studies using a target trial emulation (TTE) approach to test these potential mechanisms:

1: As simvastatin has the most potential to cross the BBB, based on its lipophilic nature and molecular weight, patients with SMI initiating simvastatin in combination with antipsychotic or mood stabilisers will have lower psychiatric hospital admissions than those initiating atorvastatin, pravastatin or rosuvastatin.

2: As simvastatin and atorvastatin inhibit p-glycoprotein and aripiprazole, risperidone and olanzapine have affinity for this:

A: patients initiating these statins in combination with these antipsychotics will have lower psychiatric hospital admissions than those initiating pravastatin (no p-glycoprotein inhibition) and the same antipsychotics.

B: patients with ongoing prescriptions of these statins initiating these antipsychotics will have lower psychiatric hospital admissions than those prescribed these statins initiating quetiapine (not a substrate of p-glycoprotein).

Through testing these hypotheses we aimed to determine the potential mechanism of action of statins in improving psychiatric symptoms in people with SMI. We use the target trial emulation framework, a causal inference approach to analysing observational data in line with a hypothetical trial, with an aim of improving study design and limiting bias (Hernan et al., 2022). This could inform the design of future RCTs and guide clinical decisions regarding the optimum combination of statins and antipsychotics in people with SMI.

## Methods

### Source population

We used the Clinical Practice Research Datalink (CPRD) Gold and Aurum databases to identify a cohort of patients diagnosed with SMI (schizophrenia, bipolar disorder and other non-organic, non-affective psychoses) between 1 January 2000 and December 2019 who were between the ages of 18 and 100 at the time of diagnosis and who received prescriptions for both antipsychotics or mood stabilisers (lithium, sodium valproate or lamotrigine), and statins. CPRD contains pseudonymised primary care data for patients across the UK and has been shown to be representative of the general UK population in respect to key patient characteristics. (Herrett et al., 2015; Wolf et al., 2019) We obtained all outcomes from the Hospital Episode Statistics (HES), a data set containing details of hospital admissions in England. As such, we limited our cohort to those patients who were eligible for linkage between CPRD and HES.

From our overall cohort, we conducted three TTEs, defined in a pre-published protocol (https://osf.io/hck8n). For each TTE we specified our hypothetical target trial to aid the design of our observational analogue (Supplementary Table 1).

Trial 1: Simvastatin vs. atorvastatin, pravastatin or rosuvastatin in people with SMI prescribed antipsychotics or mood stabilisers

### Population

We included patients from the main cohort with a diagnosis of SMI prior to statin initiation and at least one prescription for antipsychotics or mood stabilisers at therapeutic dose in the six months prior to first ever statin initiation. For antipsychotics, we first converted antipsychotic dose to olanzapine equivalent dose using the ChlorpromazineR package in R (Brown et al., 2021), using previously published conversions (Leucht et al., 2020; Leucht et al., 2016; Davis, 1974; Woods, 2003; Gardner et al., 2010). We then defined therapeutic dose as 6.5mg per day for olanzapine equivalent doses (Leucht et al., 2020). For sodium valproate, we defined therapeutic daily dose as at least 1g, based on the usual dose reported by the British National Formulary (BNF) (Joint Formulary Committee), 200mg for lithium and for lamotrigine as 200mg for monotherapy, or 100mg as adjunctive therapy with valproate as reported by the BNF.

We excluded patients whose first statin was not one of simvastatin, atorvastatin, pravastatin or rosuvastatin, those who had evidence of prior statin prescriptions (either Read codes or prescription records) or who initiated multiple statins where these belonged to both treatment and comparator arms of the TTE. We also excluded patients who had received injectable antipsychotics in the three months prior to statin initiation and those who did not have at least six months of baseline medical records (Supplementary figure 1).

### Treatment strategies, recruitment period and follow up

We assigned patients to treatment and exposure arms based on the first recorded prescription of the statins of interest. Those initiating simvastatin were in the treatment arm, and those initiating atorvastatin, pravastatin or rosuvastatin were in the comparator arm. We required that statin initiation between 1 January 2000 and 31 December 2018. Follow up ended at the earliest of the event of interest, death, end of EHR record, two years follow up, aged 100 or 1 December 2019.

### Outcomes

Our primary outcome was psychiatric admissions in the 12 months following statin initiation. Secondary outcomes were psychiatric admissions at 3, 6 and 24 months and physical health and accident and injury admissions at 3, 6, 12 and 24 months. A final secondary outcome was self-harm events. In a deviation from our published protocol, we included self-harm events reported in primary care as well as secondary care.

We retrieved hospitalisation data from HES. We defined an inpatient hospital admission as “psychiatric”, if the first episode of care had an ICD-10 “F” code as the primary diagnosis, or had a code for a mental health symptom (R41, R44, R45, R46) or observation (Z00.4, Z03.2, Z13.3, Z73) as the primary diagnosis, with an ICD-10 F code as a secondary diagnosis. We defined physical and accident and injury admissions as previously reported.(Launders et al., 2022) We defined self-harm admissions as inpatient admissions where the primary diagnosis of the first episode of care was an ICD-10 code for self-harm (Y10-34, X60-84, Y87.0), or accidents or injuries admissions, where with self-harm coded at any point during admission. We additionally included self-harm recorded in primary care using Read codes.

Trial 2A: Simvastatin or atorvastatin vs. pravastatin in people with SMI prescribed risperidone, aripiprazole or olanzapine

### Population

The population for the second trial was a subset of Trial 1 (Supplementary figure 1). Additionally, we limited this trial to patients whose first statin prescription was simvastatin, atorvastatin or pravastatin; and whose most recent antipsychotic was olanzapine, risperidone or aripiprazole at a therapeutic dose. An additional we applied a post-hoc exclusion criteria, excluding those prescribed high dose statins in the active arm as no patients were prescribed high-dose statins in the comparator arm (pravastatin).

### Treatment strategies, recruitment and follow up periods and outcomes

We assigned patients to treatment and exposure arms based on the first recorded prescription of the statins of interest. Those initiating simvastatin or atorvastatin were in the treatment arm, and those initiating pravastatin were in the comparator arm.

Recruitment and follow up periods and outcomes were as described for Trial 1.

Trial 2B: Risperidone, olanzapine and aripiprazole vs. quetiapine in patients prescribed simvastatin or atorvastatin

### Population

We included patients from the main cohort who initiated risperidone, olanzapine, aripiprazole or quetiapine and who had been prescribed simvastatin or atorvastatin in the six months prior to this.

We excluded patients who had evidence of prior prescriptions of the study antipsychotics, but not those previously prescribed other psychotropic medication. We also excluded those who were prescribed antipsychotics in both the comparator and treatment arm, those who had less than six months of baseline medical records and those who received injectable antipsychotics in the three months prior to study initiation (Supplementary figure 1).

### Treatment strategies, recruitment and follow up periods and outcomes

We assigned patients to treatment and exposure arms at one month post antipsychotic initiation to allow for titration to a therapeutic dose. Those initiating risperidone, aripiprazole or olanzapine were in the treatment arm, and those initiating quetiapine were in the comparator arm. We required patients to initiate antipsychotics between 1 January 2000 and 31 December 2018. Follow up began one month post-initiation and ended at the earliest of the event of interest, death, end of EHR record, two years follow up or 1 December 2019.

Outcomes were as described in Trial 1, with the exception that time points started at one month post initiation of antipsychotics.

### Confounding

We applied a causal framework to the identification of measured and unmeasured potential confounders (see protocol: https://osf.io/hck8n). We adjusted age, sex, deprivation and ethnicity, as well as calendar time, cardiovascular disease, prior healthcare use, specific SMI diagnosis, statin dose and duration of SMI and dyslipidaemia. We also adjusted analyses in Trial 1 for specific antipsychotic or mood stabilisers and those in Trial 2B for the time since first statin prescription.

All confounders were measured at baseline, with the exception of sex and ethnicity which were ever recorded. All were captured in primary care records, with the exception of psychiatric and self-harm hospital admissions which were taken solely from HES. We extracted ethnicity from CPRD and HES and deprivation from linked postcode data. We adjusted our multivariable models for age at study entry, year of study entry, sex, ethnicity, deprivation and the number of previous face-to-face primary care encounters in the six months before index. We grouped ethnicity into broad census categories. Where multiple ethnicities were recorded, we selected the most common, and if all equally common the most recent. We defined deprivation using quintiles of the English Index of Multiple Deprivation (IMD) 2019. We stratified baseline hazards for clustering by primary care practice.

We adjusted for the following confounders associated with mental health status: SMI diagnosis (schizophrenia, bipolar disorder or other non-organic, non-affective psychoses, not yet diagnosed); time since first SMI diagnosis and time since first antipsychotic/mood stabiliser prescription (both calculated as days and converted to years); presence of psychiatric or self-harm admissions in the year prior to index (both binary); recording of primary care self-harm events in the six months prior to index (binary); prescription of antidepressants in the six months prior to index (selective serotonin reuptake inhibitors, tricyclic antidepressants or “other”).

Finally, we adjusted for the following physical health measures: cardiovascular disease (diagnosis of hypertension, myocardial infarction or congestive heart failure); diabetes; the last recorded total cholesterol level (in the three years before index), and time since dyslipidaemia diagnosis (calculated as days and converted to years) defined by Read code or cholesterol level). Where no diagnosis of dyslipidaemia was recorded, we assumed these to be at the time of statin prescription. We also adjusted for the most recent BMI value recorded in primary care (in the five years before index, as a continuous variable), and hospital admissions for accidents and emergencies or physical health in the year prior. We adjusted for statin intensity, based on stain dosages providing less than 40% reduction in LDL cholesterol (low-moderate intensity) and those providing greater or equal to a 40% reduction (high intensity) (National Institute for Health and Care Excellence, 2023).

For trial 1 we adjusted for the specific antipsychotic or mood stabiliser prescribed, with those antipsychotics or mood stabilisers prescribed to less than 100 patients grouped as “other prescriptions”. In trial 2B we adjusted for time since first statin prescription (calculated as days and converted to years).

### Missing data

We imputed missing data for BMI value, most recent total cholesterol value and ethnicity using multiple imputation by chained equation (MICE), with coefficients from 10 imputed datasets pooled using Rubins’ rules. We included all confounders in the multivariable models in the imputation equation, as well as 2-year outcomes. We performed MICE separately for each trial, and for survival and count models. For imputation used in the Cox regression models we included the Nelson Aalen estimator. (White and Royston, 2009) We imputed missing statin dose as the median statin dose for each statin, and missing IMD as the IMD of the primary care practice.

### Analysis

We described patient characteristics for each trial. In our primary analysis we used Cox regression to quantify the hazard ratio (HR) of all outcomes between trial arms. We tested the proportional hazard assumption using Schoenfeld residuals and plotted Kaplan Meier plots of unadjusted models. We then adjusted all models for the previously described confounders. We tested for interactions between trial arms and SMI subtype for 12-month outcomes in the cox regression models for all three trials, and where we found significant interactions we present stratified results by SMI diagnosis. Our secondary analysis utilised negative binomial regression to assess the incidence rate ratio (IRR) of outcome events between the arms of each trial. We present all HRs and IRRs with 95% confidence intervals (95%CI). We assessed all outcomes using an observational analogue of intention-to-treat analysis, with patients analysed in their original treatment arm assignments.

### Sensitivity analyses

We defined sensitivity analyses a priori. We investigated the per-protocol effect for the primary outcome of each trial and the impact of using inverse probability weighting to control for confounders (See supplementary methods). Finally, we had prespecified a sensitivity analysis to widen the inclusion criteria of trial 1 and 2 to include patients who had used statins previously (https://osf.io/hck8n), but with a six-month washout period of no statin use, prior to statin reinitiation. However, as this method only yielded a 5 and 6% increase in sample size respectively compared to the more stringent design, and only yielded an extra 19 patients in the smallest arm of trial 2A, we did not perform this analysis.

## Results

We identified 72,096 adults in CPRD with a diagnosis of SMI who were ever prescribed both statins and antipsychotics or mood stabilisers, were registered with a primary care practice at any point from 1 January 2000 to 31 December 2019 and eligible for HES linkage. Of these, 19,033 were eligible for Trial 1, 6,081 for Trial 2A and 9,398 for Trial 2B (Supplementary figure 1).

Trial 1: Simvastatin vs. atorvastatin, pravastatin or rosuvastatin in people with SMI prescribed antipsychotics or mood stabilisers

The proportional hazard assumption held for all four 12-month outcomes (Supplementary Figure 2). We found no evidence of interaction effects between SMI diagnosis and trial arm for any 12-month outcomes.

### Primary outcome: Psychiatric hospital admissions at 12 months

Contrary to our hypothesis, patients initiating simvastatin were more likely to have a psychiatric admission in the 12 months following initiation than those initiating atorvastatin, pravastatin or rosuvastatin in unadjusted analysis (HR: 1.22; 95%CI: 1.07-1.39). However, this did not persist following adjustment (adjusted HR: 1.07; 95%CI:0.88-1.31, Table 2; Figure 1).

**Figure 1:**
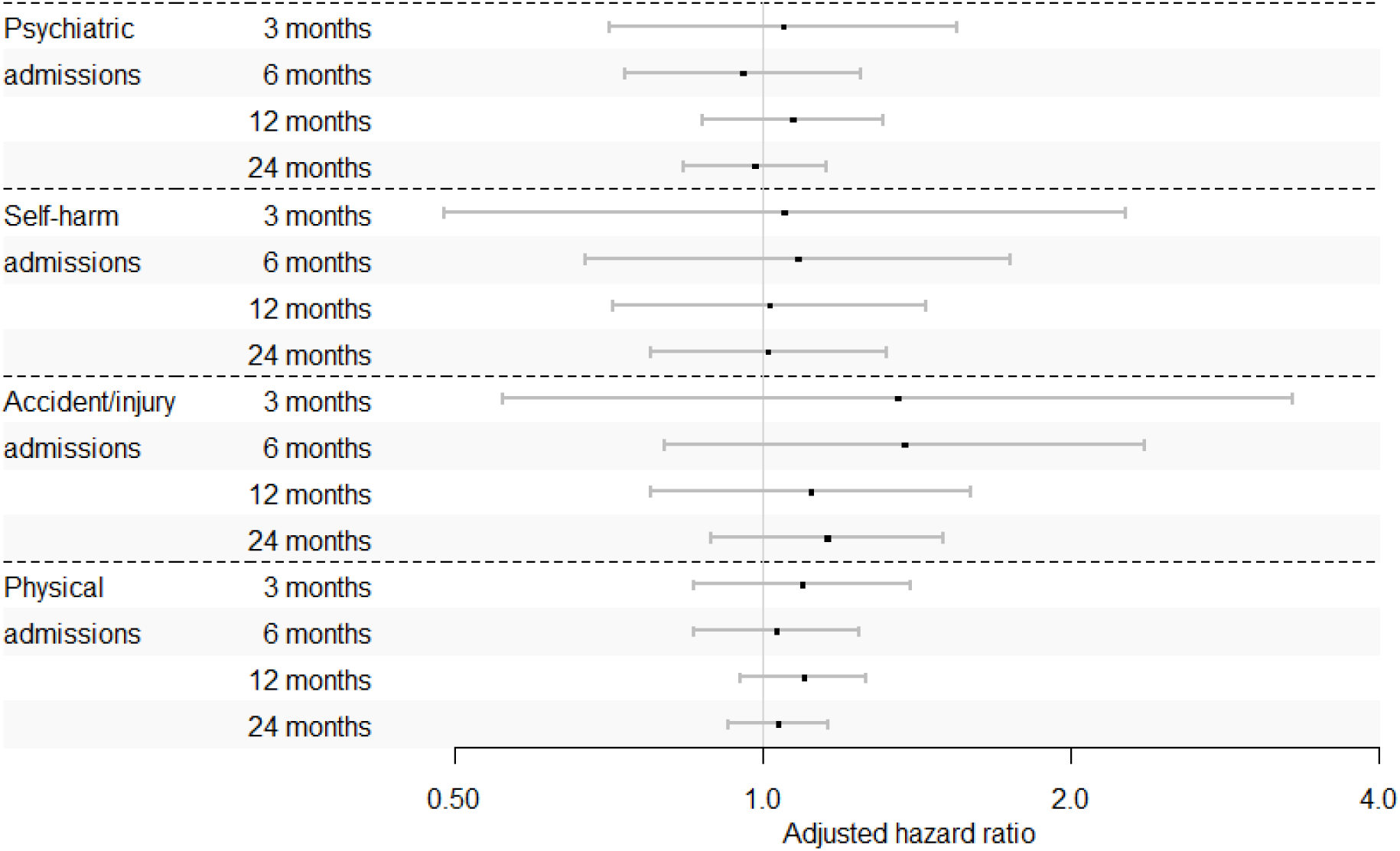
Cox regression results for trial 1: Simvastatin vs. atorvastatin, pravastatin or rosuvastatin in people with SMI prescribed antipsychotics or mood stabilisers.

**Table 1:**
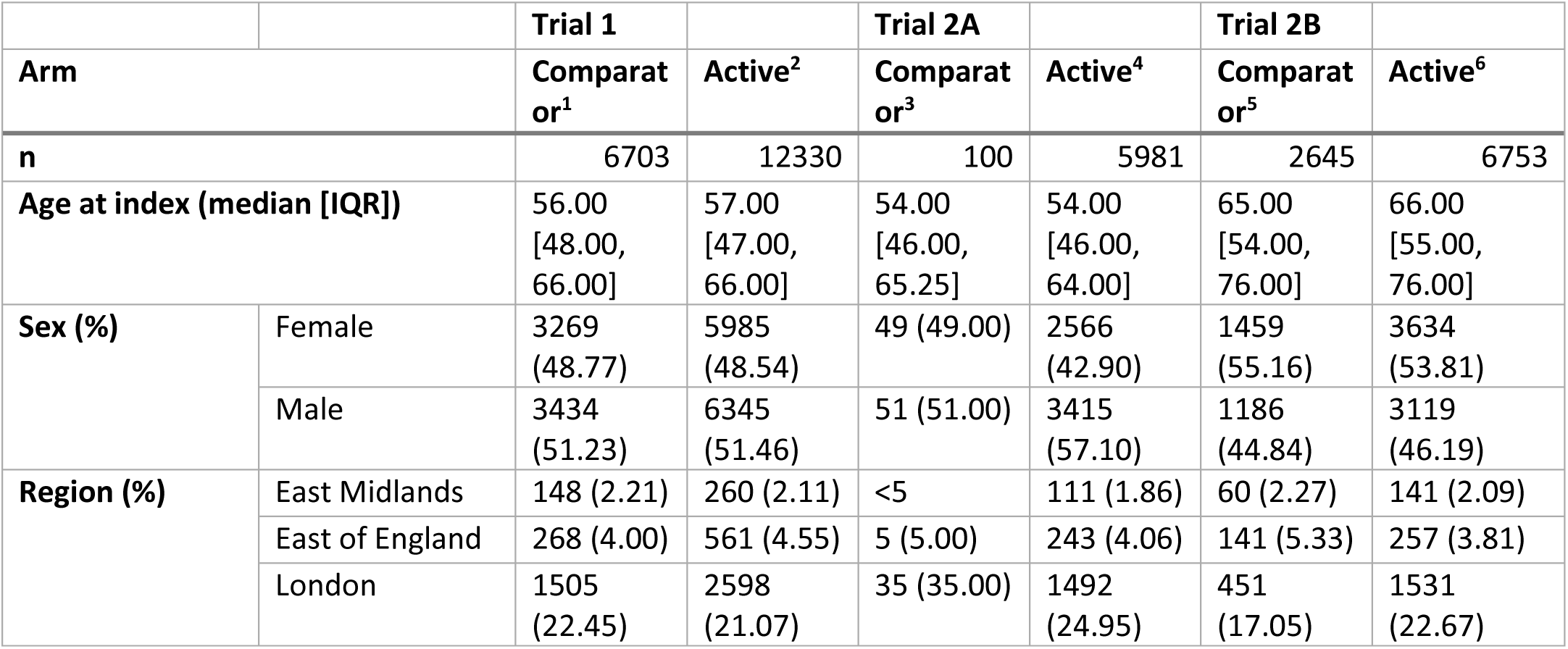
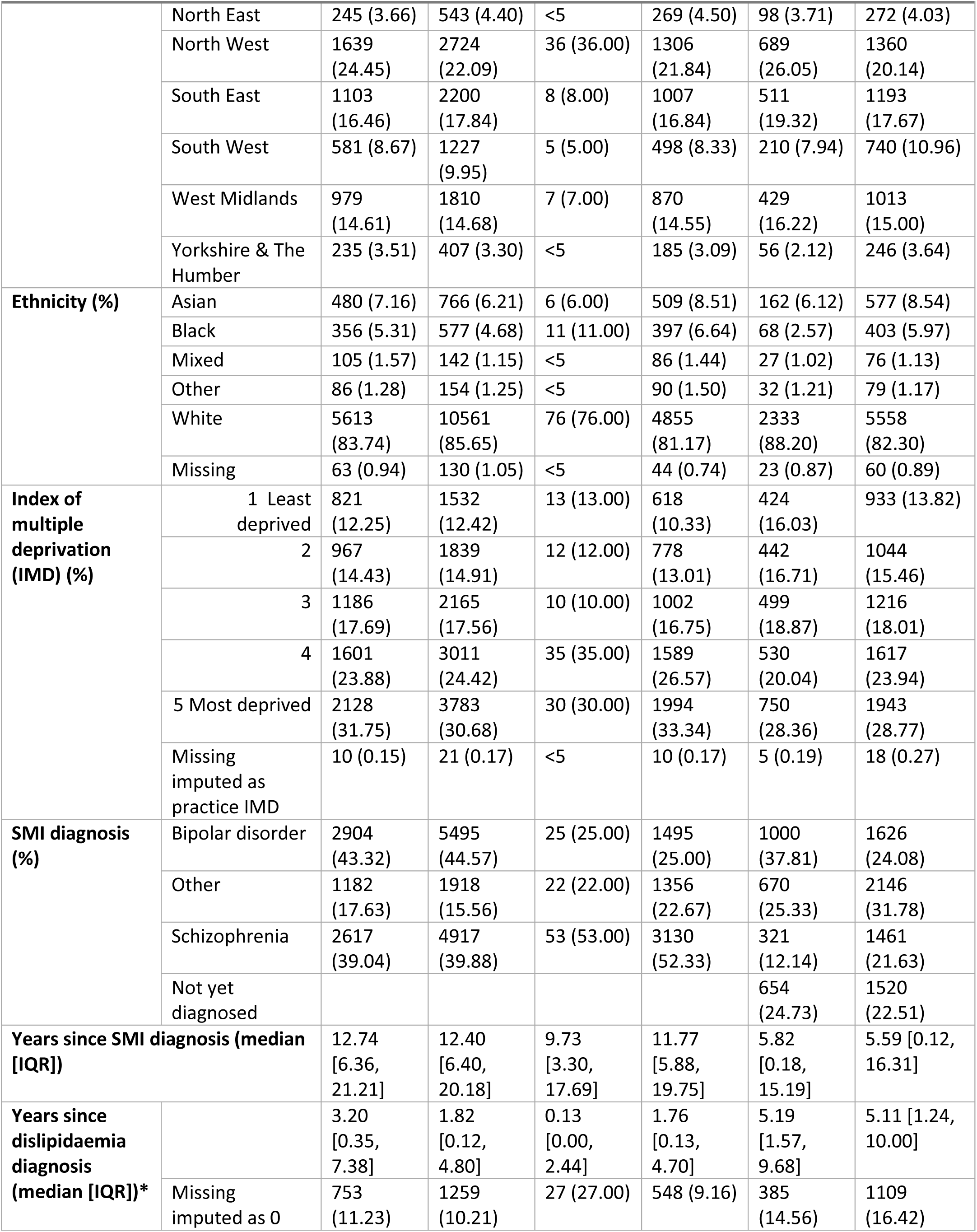
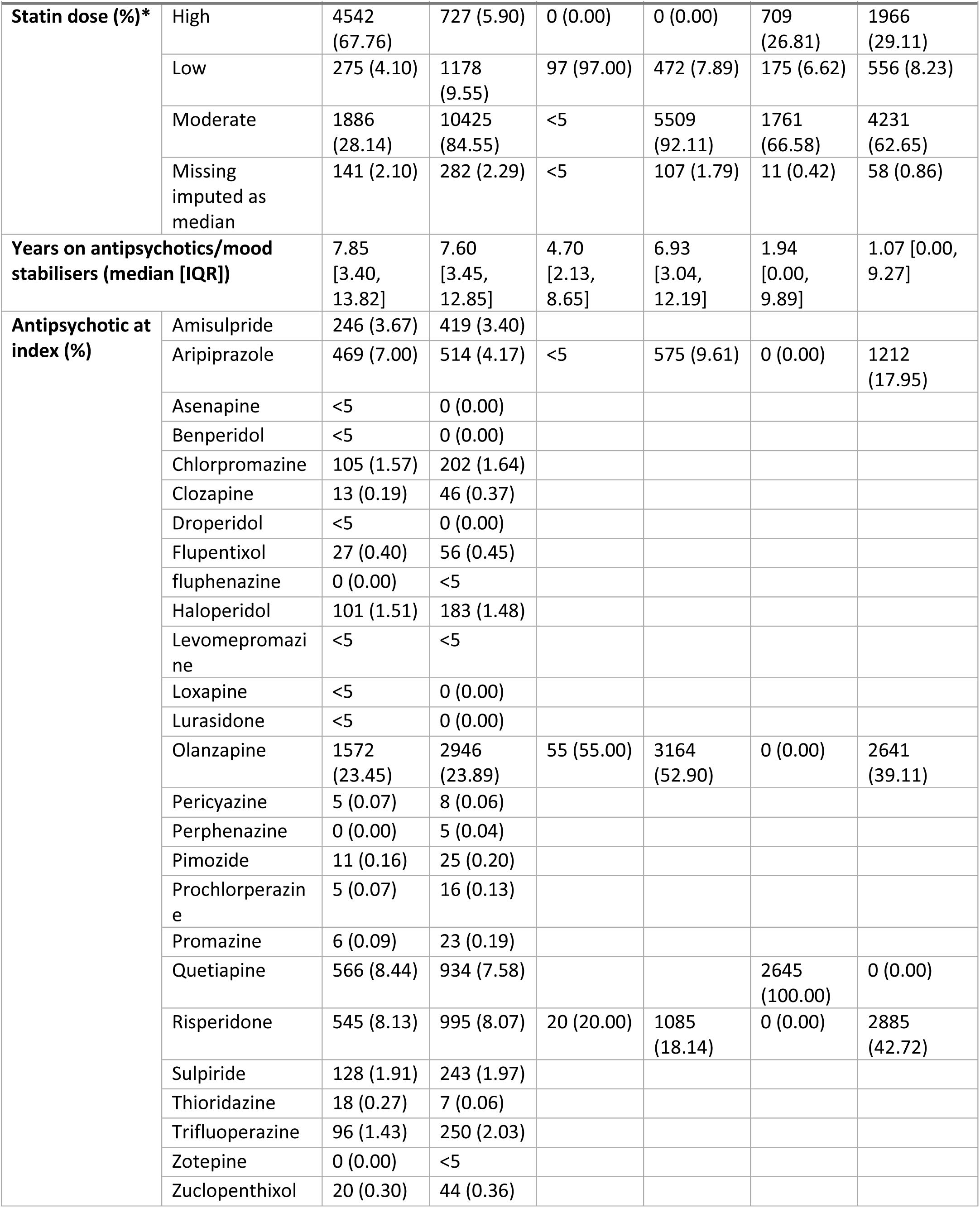
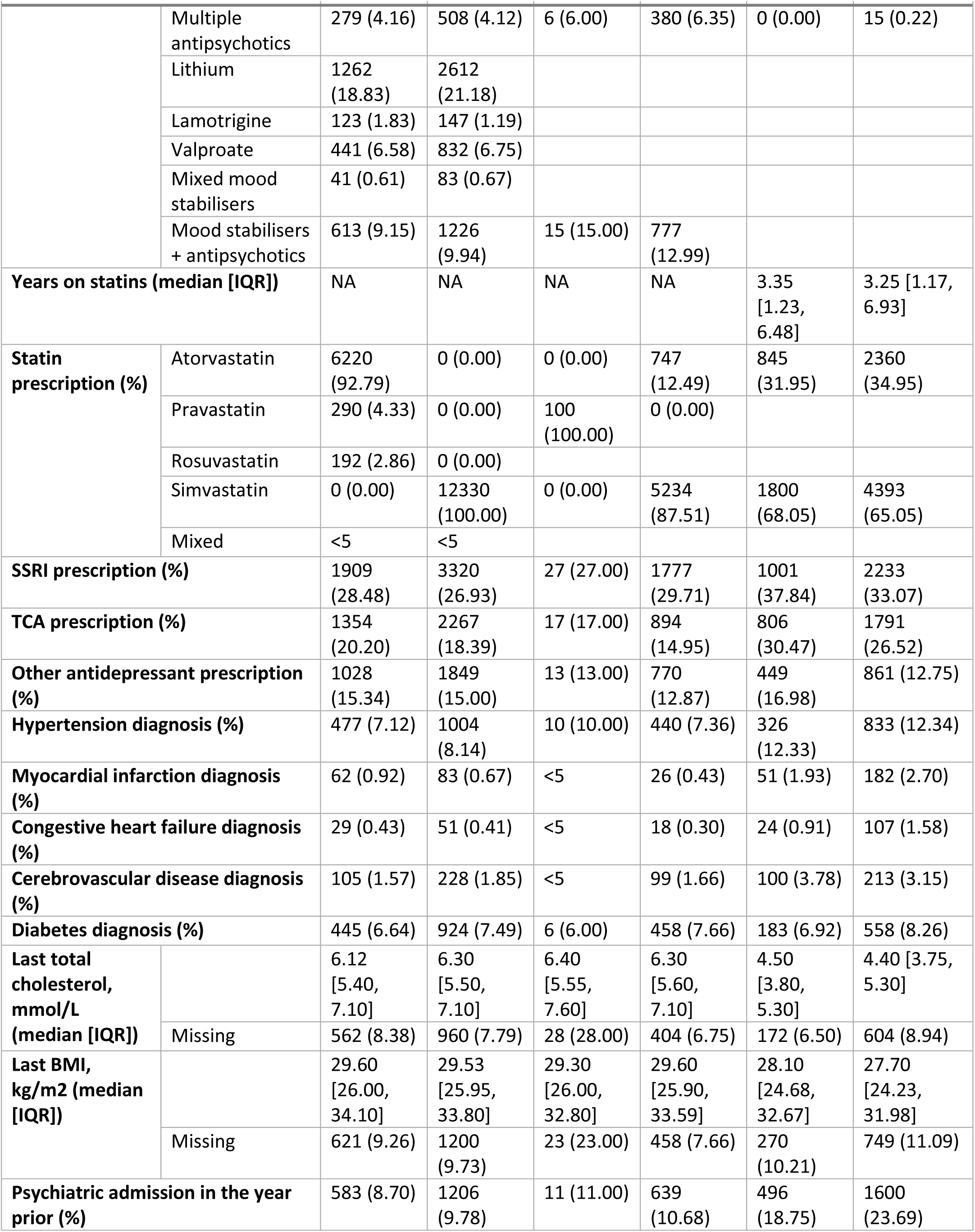
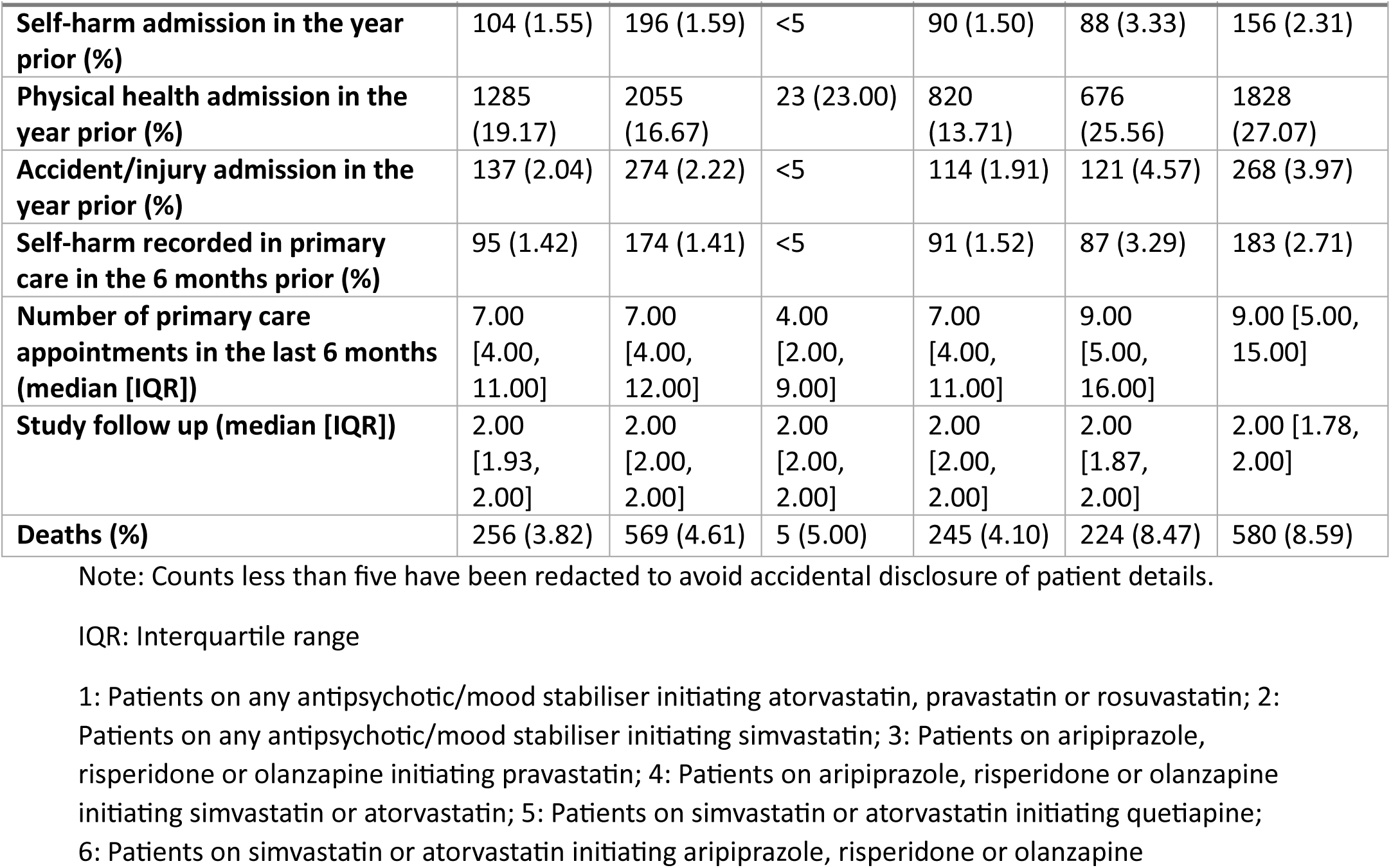
Patient characteristics for three target trial emulation studies.

|  |  | **Trial 1** |  | **Trial 2A** |  | **Trial 2B** |  |
| --- | --- | --- | --- | --- | --- | --- | --- |
| **Arm** |  | **Comparat**<br>**or<sup>1</sup>** | **Active<sup>2</sup>** | **Comparat**<br>**or<sup>3</sup>** | **Active<sup>4</sup>** | **Comparat**<br>**or<sup>5</sup>** | **Active<sup>6</sup>** |
| **n** |  | 6703 | 12330 | 100 | 5981 | 2645 | 6753 |
| **Age at index (median [IQR])** |  | 56.00<br>[48.00,<br>66.00] | 57.00<br>[47.00,<br>66.00] | 54.00<br>[46.00,<br>65.25] | 54.00<br>[46.00,<br>64.00] | 65.00<br>[54.00,<br>76.00] | 66.00<br>[55.00,<br>76.00] |
| **Sex (%)** | Female | 3269<br>(48.77) | 5985<br>(48.54) | 49 (49.00) | 2566<br>(42.90) | 1459<br>(55.16) | 3634<br>(53.81) |
|  | Male | 3434<br>(51.23) | 6345<br>(51.46) | 51 (51.00) | 3415<br>(57.10) | 1186<br>(44.84) | 3119<br>(46.19) |
| **Region (%)** | East Midlands | 148 (2.21) | 260 (2.11) | <5 | 111 (1.86) | 60 (2.27) | 141 (2.09) |
|  | East of England | 268 (4.00) | 561 (4.55) | 5 (5.00) | 243 (4.06) | 141 (5.33) | 257 (3.81) |
|  | London | 1505<br>(22.45) | 2598<br>(21.07) | 35 (35.00) | 1492<br>(24.95) | 451<br>(17.05) | 1531<br>(22.67) |
| **Arm** |  | **Comparat**<br>**or<sup>1</sup>** | **Active<sup>2</sup>** | **Comparat**<br>**or<sup>3</sup>** | **Active<sup>4</sup>** | **Comparat**<br>**or<sup>5</sup>** | **Active<sup>6</sup>** |
|  | North East | 245 (3.66) | 543 (4.40) | <5 | 269 (4.50) | 98 (3.71) | 272 (4.03) |
|  | North West | 1639<br>(24.45) | 2724<br>(22.09) | 36 (36.00) | 1306<br>(21.84) | 689<br>(26.05) | 1360<br>(20.14) |
|  | South East | 1103<br>(16.46) | 2200<br>(17.84) | 8 (8.00) | 1007<br>(16.84) | 511<br>(19.32) | 1193<br>(17.67) |
|  | South West | 581 (8.67) | 1227<br>(9.95) | 5 (5.00) | 498 (8.33) | 210 (7.94) | 740 (10.96) |
|  | West Midlands | 979<br>(14.61) | 1810<br>(14.68) | 7 (7.00) | 870<br>(14.55) | 429<br>(16.22) | 1013<br>(15.00) |
|  | Yorkshire & The<br>Humber | 235 (3.51) | 407 (3.30) | <5 | 185 (3.09) | 56 (2.12) | 246 (3.64) |
| **Ethnicity (%)** | Asian | 480 (7.16) | 766 (6.21) | 6 (6.00) | 509 (8.51) | 162 (6.12) | 577 (8.54) |
|  | Black | 356 (5.31) | 577 (4.68) | 11 (11.00) | 397 (6.64) | 68 (2.57) | 403 (5.97) |
|  | Mixed | 105 (1.57) | 142 (1.15) | <5 | 86 (1.44) | 27 (1.02) | 76 (1.13) |
|  | Other | 86 (1.28) | 154 (1.25) | <5 | 90 (1.50) | 32 (1.21) | 79 (1.17) |
|  | White | 5613<br>(83.74) | 10561<br>(85.65) | 76 (76.00) | 4855<br>(81.17) | 2333<br>(88.20) | 5558<br>(82.30) |
|  | Missing | 63 (0.94) | 130 (1.05) | <5 | 44 (0.74) | 23 (0.87) | 60 (0.89) |
| **Index of**<br>**multiple**<br>**deprivation**<br>**(IMD) (%)** | 1 Least<br>deprived | 821<br>(12.25) | 1532<br>(12.42) | 13 (13.00) | 618<br>(10.33) | 424<br>(16.03) | 933 (13.82) |
|  | 2 | 967<br>(14.43) | 1839<br>(14.91) | 12 (12.00) | 778<br>(13.01) | 442<br>(16.71) | 1044<br>(15.46) |
|  | 3 | 1186<br>(17.69) | 2165<br>(17.56) | 10 (10.00) | 1002<br>(16.75) | 499<br>(18.87) | 1216<br>(18.01) |
|  | 4 | 1601<br>(23.88) | 3011<br>(24.42) | 35 (35.00) | 1589<br>(26.57) | 530<br>(20.04) | 1617<br>(23.94) |
|  | 5 Most deprived | 2128<br>(31.75) | 3783<br>(30.68) | 30 (30.00) | 1994<br>(33.34) | 750<br>(28.36) | 1943<br>(28.77) |
|  | Missing<br>imputed as<br>practice IMD | 10 (0.15) | 21 (0.17) | <5 | 10 (0.17) | 5 (0.19) | 18 (0.27) |
| **SMI diagnosis**<br>**(%)** | Bipolar disorder | 2904<br>(43.32) | 5495<br>(44.57) | 25 (25.00) | 1495<br>(25.00) | 1000<br>(37.81) | 1626<br>(24.08) |
|  | Other | 1182<br>(17.63) | 1918<br>(15.56) | 22 (22.00) | 1356<br>(22.67) | 670<br>(25.33) | 2146<br>(31.78) |
|  | Schizophrenia | 2617<br>(39.04) | 4917<br>(39.88) | 53 (53.00) | 3130<br>(52.33) | 321<br>(12.14) | 1461<br>(21.63) |
|  | Not yet<br>diagnosed |  |  |  |  | 654<br>(24.73) | 1520<br>(22.51) |
| **Years since SMI diagnosis (median**<br>**[IQR])** |  | 12.74<br>[6.36,<br>21.21] | 12.40<br>[6.40,<br>20.18] | 9.73<br>[3.30,<br>17.69] | 11.77<br>[5.88,<br>19.75] | 5.82<br>[0.18,<br>15.19] | 5.59 [0.12,<br>16.31] |
| **Years since**<br>**dislipidaemia**<br>**diagnosis**<br>**(median [IQR])\*** |  | 3.20<br>[0.35,<br>7.38] | 1.82<br>[0.12,<br>4.80] | 0.13<br>[0.00,<br>2.44] | 1.76<br>[0.13,<br>4.70] | 5.19<br>[1.57,<br>9.68] | 5.11 [1.24,<br>10.00] |
|  | Missing<br>imputed as 0 | 753<br>(11.23) | 1259<br>(10.21) | 27 (27.00) | 548 (9.16) | 385<br>(14.56) | 1109<br>(16.42) |

|  |  | Trial 1 |  | Trial 2A |  | Trial 2B |  |
| --- | --- | --- | --- | --- | --- | --- | --- |
| Arm |  | Comparator<sup>1</sup> | Active<sup>2</sup> | Comparator<sup>3</sup> | Active<sup>4</sup> | Comparator<sup>5</sup> | Active<sup>6</sup> |
| Statin dose (%)* | High | 4542 (67.76) | 727 (5.90) | 0 (0.00) | 0 (0.00) | 709 (26.81) | 1966 (29.11) |
|  | Low | 275 (4.10) | 1178 (9.55) | 97 (97.00) | 472 (7.89) | 175 (6.62) | 556 (8.23) |
|  | Moderate | 1886 (28.14) | 10425 (84.55) | <5 | 5509 (92.11) | 1761 (66.58) | 4231 (62.65) |
|  | Missing imputed as median | 141 (2.10) | 282 (2.29) | <5 | 107 (1.79) | 11 (0.42) | 58 (0.86) |
| Years on antipsychotics/mood stabilisers (median [IQR]) |  | 7.85 [3.40, 13.82] | 7.60 [3.45, 12.85] | 4.70 [2.13, 8.65] | 6.93 [3.04, 12.19] | 1.94 [0.00, 9.89] | 1.07 [0.00, 9.27] |
| Antipsychotic at index (%) | Amisulpride | 246 (3.67) | 419 (3.40) |  |  |  |  |
|  | Aripiprazole | 469 (7.00) | 514 (4.17) | <5 | 575 (9.61) | 0 (0.00) | 1212 (17.95) |
|  | Asenapine | <5 | 0 (0.00) |  |  |  |  |
|  | Benperidol | <5 | 0 (0.00) |  |  |  |  |
|  | Chlorpromazine | 105 (1.57) | 202 (1.64) |  |  |  |  |
|  | Clozapine | 13 (0.19) | 46 (0.37) |  |  |  |  |
|  | Droperidol | <5 | 0 (0.00) |  |  |  |  |
|  | Flupentixol | 27 (0.40) | 56 (0.45) |  |  |  |  |
|  | fluphenazine | 0 (0.00) | <5 |  |  |  |  |
|  | Haloperidol | 101 (1.51) | 183 (1.48) |  |  |  |  |
|  | Levomepromazine | <5 | <5 |  |  |  |  |
|  | Loxapine | <5 | 0 (0.00) |  |  |  |  |
|  | Lurasidone | <5 | 0 (0.00) |  |  |  |  |
|  | Olanzapine | 1572 (23.45) | 2946 (23.89) | 55 (55.00) | 3164 (52.90) | 0 (0.00) | 2641 (39.11) |
|  | Pericyazine | 5 (0.07) | 8 (0.06) |  |  |  |  |
|  | Perphenazine | 0 (0.00) | 5 (0.04) |  |  |  |  |
|  | Pimozide | 11 (0.16) | 25 (0.20) |  |  |  |  |
|  | Prochlorperazine | 5 (0.07) | 16 (0.13) |  |  |  |  |
|  | Promazine | 6 (0.09) | 23 (0.19) |  |  |  |  |
|  | Quetiapine | 566 (8.44) | 934 (7.58) |  |  | 2645 (100.00) | 0 (0.00) |
|  | Risperidone | 545 (8.13) | 995 (8.07) | 20 (20.00) | 1085 (18.14) | 0 (0.00) | 2885 (42.72) |
|  | Sulpiride | 128 (1.91) | 243 (1.97) |  |  |  |  |
|  | Thioridazine | 18 (0.27) | 7 (0.06) |  |  |  |  |
|  | Trifluoperazine | 96 (1.43) | 250 (2.03) |  |  |  |  |
|  | Zotepine | 0 (0.00) | <5 |  |  |  |  |
|  | Zuclopenthixol | 20 (0.30) | 44 (0.36) |  |  |  |  |
| **Arm** |  | **Comparator<sup>1</sup>** | **Active<sup>2</sup>** | **Comparator<sup>3</sup>** | **Active<sup>4</sup>** | **Comparator<sup>5</sup>** | **Active<sup>6</sup>** |
|  | Multiple antipsychotics | 279 (4.16) | 508 (4.12) | 6 (6.00) | 380 (6.35) | 0 (0.00) | 15 (0.22) |
|  | Lithium | 1262 (18.83) | 2612 (21.18) |  |  |  |  |
|  | Lamotrigine | 123 (1.83) | 147 (1.19) |  |  |  |  |
|  | Valproate | 441 (6.58) | 832 (6.75) |  |  |  |  |
|  | Mixed mood stabilisers | 41 (0.61) | 83 (0.67) |  |  |  |  |
|  | Mood stabilisers + antipsychotics | 613 (9.15) | 1226 (9.94) | 15 (15.00) | 777 (12.99) |  |  |
| **Years on statins (median [IQR])** |  | NA | NA | NA | NA | 3.35 [1.23, 6.48] | 3.25 [1.17, 6.93] |
| **Statin prescription (%)** | Atorvastatin | 6220 (92.79) | 0 (0.00) | 0 (0.00) | 747 (12.49) | 845 (31.95) | 2360 (34.95) |
|  | Pravastatin | 290 (4.33) | 0 (0.00) | 100 (100.00) | 0 (0.00) |  |  |
|  | Rosuvastatin | 192 (2.86) | 0 (0.00) |  |  |  |  |
|  | Simvastatin | 0 (0.00) | 12330 (100.00) | 0 (0.00) | 5234 (87.51) | 1800 (68.05) | 4393 (65.05) |
|  | Mixed | <5 | <5 |  |  |  |  |
| **SSRI prescription (%)** |  | 1909 (28.48) | 3320 (26.93) | 27 (27.00) | 1777 (29.71) | 1001 (37.84) | 2233 (33.07) |
| **TCA prescription (%)** |  | 1354 (20.20) | 2267 (18.39) | 17 (17.00) | 894 (14.95) | 806 (30.47) | 1791 (26.52) |
| **Other antidepressant prescription (%)** |  | 1028 (15.34) | 1849 (15.00) | 13 (13.00) | 770 (12.87) | 449 (16.98) | 861 (12.75) |
| **Hypertension diagnosis (%)** |  | 477 (7.12) | 1004 (8.14) | 10 (10.00) | 440 (7.36) | 326 (12.33) | 833 (12.34) |
| **Myocardial infarction diagnosis (%)** |  | 62 (0.92) | 83 (0.67) | <5 | 26 (0.43) | 51 (1.93) | 182 (2.70) |
| **Congestive heart failure diagnosis (%)** |  | 29 (0.43) | 51 (0.41) | <5 | 18 (0.30) | 24 (0.91) | 107 (1.58) |
| **Cerebrovascular disease diagnosis (%)** |  | 105 (1.57) | 228 (1.85) | <5 | 99 (1.66) | 100 (3.78) | 213 (3.15) |
| **Diabetes diagnosis (%)** |  | 445 (6.64) | 924 (7.49) | 6 (6.00) | 458 (7.66) | 183 (6.92) | 558 (8.26) |
| **Last total cholesterol, mmol/L (median [IQR])** |  | 6.12 [5.40, 7.10] | 6.30 [5.50, 7.10] | 6.40 [5.55, 7.60] | 6.30 [5.60, 7.10] | 4.50 [3.80, 5.30] | 4.40 [3.75, 5.30] |
|  | Missing | 562 (8.38) | 960 (7.79) | 28 (28.00) | 404 (6.75) | 172 (6.50) | 604 (8.94) |
| **Last BMI, kg/m2 (median [IQR])** |  | 29.60 [26.00, 34.10] | 29.53 [25.95, 33.80] | 29.30 [26.00, 32.80] | 29.60 [25.90, 33.59] | 28.10 [24.68, 32.67] | 27.70 [24.23, 31.98] |
|  | Missing | 621 (9.26) | 1200 (9.73) | 23 (23.00) | 458 (7.66) | 270 (10.21) | 749 (11.09) |
| **Psychiatric admission in the year prior (%)** |  | 583 (8.70) | 1206 (9.78) | 11 (11.00) | 639 (10.68) | 496 (18.75) | 1600 (23.69) |
| **Arm** |  | **Comparat**<br>**or<sup>1</sup>** | **Active<sup>2</sup>** | **Comparat**<br>**or<sup>3</sup>** | **Active<sup>4</sup>** | **Comparat**<br>**or<sup>5</sup>** | **Active<sup>6</sup>** |
| **Self-harm admission in the year prior (%)** |  | 104 (1.55) | 196 (1.59) | <5 | 90 (1.50) | 88 (3.33) | 156 (2.31) |
| **Physical health admission in the year prior (%)** |  | 1285 (19.17) | 2055 (16.67) | 23 (23.00) | 820 (13.71) | 676 (25.56) | 1828 (27.07) |
| **Accident/injury admission in the year prior (%)** |  | 137 (2.04) | 274 (2.22) | <5 | 114 (1.91) | 121 (4.57) | 268 (3.97) |
| **Self-harm recorded in primary care in the 6 months prior (%)** |  | 95 (1.42) | 174 (1.41) | <5 | 91 (1.52) | 87 (3.29) | 183 (2.71) |
| **Number of primary care appointments in the last 6 months (median [IQR])** |  | 7.00 [4.00, 11.00] | 7.00 [4.00, 12.00] | 4.00 [2.00, 9.00] | 7.00 [4.00, 11.00] | 9.00 [5.00, 16.00] | 9.00 [5.00, 15.00] |
| **Study follow up (median [IQR])** |  | 2.00 [1.93, 2.00] | 2.00 [2.00, 2.00] | 2.00 [2.00, 2.00] | 2.00 [2.00, 2.00] | 2.00 [1.87, 2.00] | 2.00 [1.78, 2.00] |
| **Deaths (%)** |  | 256 (3.82) | 569 (4.61) | 5 (5.00) | 245 (4.10) | 224 (8.47) | 580 (8.59) |

Note: Counts less than five have been redacted to avoid accidental disclosure of patient details.
IQR: Interquartile range
1: Patients on any antipsychotic/mood stabiliser initiating atorvastatin, pravastatin or rosuvastatin; 2: Patients on any antipsychotic/mood stabiliser initiating simvastatin; 3: Patients on aripiprazole, risperidone or olanzapine initiating pravastatin; 4: Patients on aripiprazole, risperidone or olanzapine initiating simvastatin or atorvastatin; 5: Patients on simvastatin or atorvastatin initiating quetiapine; 6: Patients on simvastatin or atorvastatin initiating aripiprazole, risperidone or olanzapine

**Table 2:** Unadjusted and adjusted Cox regression results for the three target trial emulation studies.

|  |  | 3 months, hazard ratio (95% CI) | 6 months, hazard ratio (95% CI) | 12 months, hazard ratio (95% CI) | 24 months, hazard ratio (95% CI) |
| --- | --- | --- | --- | --- | --- |
| **TRIAL 1: Simvastatin vs. atorvastatin, pravastatin or rosuvastatin in people with SMI prescribed antipsychotics or mood stabilisers** |  |  |  |  |  |
| Psychiatric admissions | Unadjusted | 1.42 (1.11-1.82) | 1.27 (1.07-1.50) | 1.22 (1.07-1.39) | 1.16 (1.04-1.29) |
|  | Adjusted | 1.05 (0.71-1.55) | 0.96 (0.73-1.25) | 1.07 (0.88-1.31) | 0.98 (0.84-1.16) |
| Self-harm events | Unadjusted | 1.11 (0.75-1.63) | 1.01 (0.77-1.31) | 1.06 (0.86-1.30) | 1.04 (0.88-1.22) |
|  | Adjusted | 1.05 (0.49-2.26) | 1.08 (0.67-1.75) | 1.02 (0.72-1.44) | 1.01 (0.78-1.32) |
| Physical health admissions | Unadjusted | 0.93 (0.80-1.08) | 0.95 (0.84-1.06) | 0.98 (0.90-1.07) | 0.95 (0.89-1.03) |
|  | Adjusted | 1.09 (0.86-1.40) | 1.03 (0.86-1.24) | 1.10 (0.95-1.26) | 1.04 (0.93-1.16) |
| Accident/injury admissions | Unadjusted | 1.00 (0.64-1.59) | 1.07 (0.79-1.44) | 1.04 (0.84-1.29) | 1.10 (0.93-1.29) |
|  | Adjusted | 1.36 (0.56-3.30) | 1.38 (0.80-2.36) | 1.12 (0.78-1.60) | 1.16 (0.89-1.50) |
| **TRIAL 2A: Simvastatin or atorvastatin vs. pravastatin in people with SMI prescribed risperidone, aripiprazole or olanzapine** |  |  |  |  |  |
| Psychiatric admissions | Unadjusted | 0.89 (0.16-4.80) | 0.70 (0.24-2.07) | 0.77 (0.32-1.87) | 0.87 (0.43-1.75) |
|  | Adjusted | 0.46 (0.05-4.70) | 0.49 (0.13-1.92) | 0.77 (0.28-2.15) | 0.93 (0.41-2.10) |
| Self-harm events | Unadjusted | -- | 0.68 (0.07-7.09) | 1.47 (0.17-12.84) | 1.64 (0.36-7.42) |
|  | Adjusted | -- | 0.10 (0.00-34.41) | 1.32 (0.02-71.14) | 2.19 (0.32-14.79) |
| Physical health admissions | Unadjusted | 0.93 (0.25-3.45) | 0.74 (0.29-1.94) | 0.58 (0.29-1.16) | 0.65 (0.38-1.11) |
|  | Adjusted | 1.11 (0.22-5.65) | 0.63 (0.20- 1.93) | 0.63 (0.27-1.45) | 0.61 (0.33-1.14) |
| Accident/injury admissions | Unadjusted | -- | -- | 0.99 (0.12-8.35) | 0.87 (0.18-4.09) |
|  | Adjusted | -- | -- | 3.02 (0.16-57.75) | 1.28 (0.21-7.78) |
| **TRIAL 2B: Risperidone, olanzapine and aripiprazole vs. quetiapine in patients prescribed simvastatin or atorvastatin** |  |  |  |  |  |
| Psychiatric admissions | Unadjusted | 1.04 (0.82-1.32) | 1.07 (0.89-1.28) | 0.97 (0.83-1.12) | 0.99 (0.87-1.13) |
|  | Adjusted | 0.99 (0.76-1.28) | 1.05 (0.87-1.28) | 0.93 (0.79-1.09) | 0.96 (0.84-1.10) |
| Self-harm events | Unadjusted | 0.78 (0.50-1.21) | 0.78 (0.56-1.09) | 0.73 (0.56-0.96) | 0.72 (0.57-0.91) |
|  | Adjusted | 0.79 (0.43-1.46) | 0.90 (0.60-1.35) | 0.80 (0.58-1.09) | 0.82 (0.63-1.09) |
| Physical health admissions | Unadjusted | 1.06 (0.88-1.27) | 1.03 (0.90-1.18) | 0.97 (0.87-1.08) | 0.96 (0.87-1.05) |
|  | Adjusted | 1.01 (0.83-1.23) | 1.00 (0.87-1.16) | 0.93 (0.83-1.05) | 0.92 (0.83-1.01) |
| Accident/injury admissions | Unadjusted | 1.26 (0.79-2.00) | 1.18 (0.84-1.67) | 0.89 (0.70-1.14) | 0.89 (0.73-1.09) |
|  | Adjusted | 1.30 (0.77-2.20) | 1.13 (0.77-1.65) | 0.87 (0.67-1.13) | 0.87 (0.71-1.08) |

### Secondary outcomes

The psychiatric admission rate was higher in those prescribed simvastatin than in the comparator arm at 3, 6 and 24 months in the unadjusted analysis, however in adjusted analysis we did not observe a difference in admission rate (Table 2). We found no evidence of a difference in self-harm events, accident and injury or physical health admissions at 3, 6, 12 or 24 months in unadjusted or adjusted analyses (Table 2; Figure 1).

### Negative binomial models and sensitivity analyses

The adjusted analysis using negative binomial models largely agreed with that of the Cox regression analysis. The exception was self-harm events at 3 months which were elevated in those prescribed simvastatin compared to those prescribed atorvastatin, pravastatin or rosuvastatin (IRR: 1.95; 95%CI:1.03-3.70; Supplementary table 2). The IPW model found higher rates of psychiatric admissions and self-harm events at 3 months in those prescribed simvastatin and more accident/injury admissions at 6 months (Supplementary table 3). As statin dose remained unbalanced after IPW (see supplementary methods), we re-ran the IPW-weighted regression additionally adjusting for this, however this did not change the results. The per-protocol analysis agreed with the main results (Supplementary table 4).

Trial 2A: Simvastatin or atorvastatin vs. pravastatin in people with SMI prescribed risperidone, aripiprazole or olanzapine

The proportional hazard assumption held for all four outcomes at 12 months (Supplementary Figure 3). Small numbers in the comparator arm (n=100) meant that we were unable to reliably test for interactions between trial arm and SMI diagnosis.

### Primary outcome: Psychiatric hospital admissions at 12 months

There was no evidence of a difference in psychiatric admissions at 12 months between patients prescribed simvastatin or atorvastatin and those prescribed pravastatin in unadjusted (HR: 0.77, 95%CI:0.32-1.87) or adjusted models (HR: 0.77; 95%CI:0.28-2.15, Table 2, Figure 2). However, in both these analyses we found a large negative effect size with wide confidence intervals.

**Figure 2:**
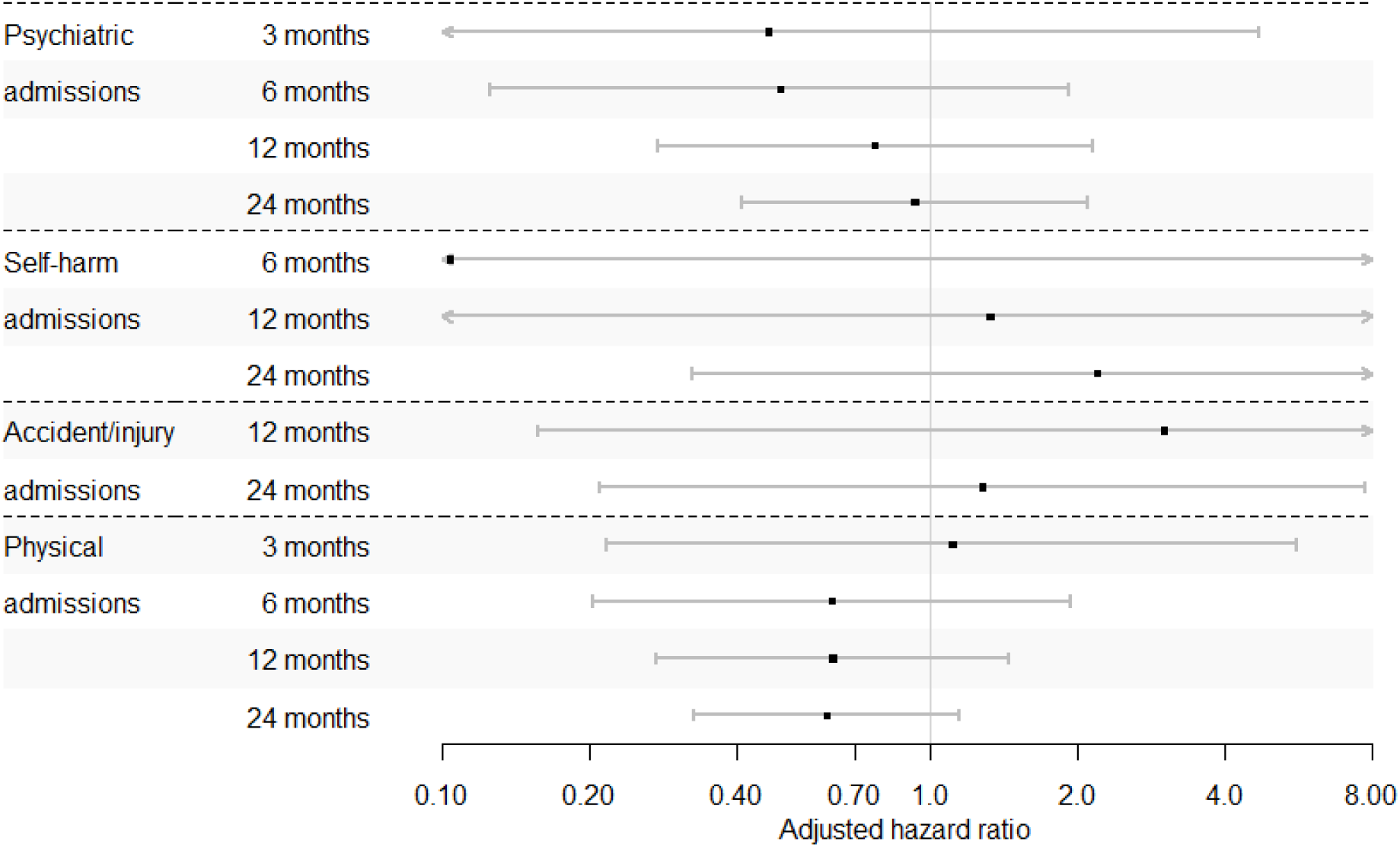
Cox regression results for trial 2A: Simvastatin or atorvastatin vs. pravastatin in people with SMI prescribed risperidone, aripiprazole or olanzapine.

### Secondary outcomes

We observed no difference in psychiatric, self-harm events, accident and injury or physical health admissions at any time point (Table 2, Figure 2). We were unable to model accident/injury admissions or self-harm admissions at 3 months and accident/injury admissions at 6 months due to the low number of outcome events.

### Negative binomial models and sensitivity analyses

The adjusted analysis using negative binomial models agreed with that of the Cox regression analysis for all outcomes (Supplementary table 2). The IPW model found a higher rate of physical health admissions at 3, 6 and 12 months in those treated with simvastatin or atorvastatin (Supplementary table 3). However, the confidence intervals were very wide and the IPW failed to balance covariates (see supplementary methods) resulting in a very small effective population size. The per-protocol analysis agreed with the main analysis, but small sample size and reduced follow p time resulted in large confidence intervals.

Trial 2B: Risperidone, olanzapine and aripiprazole vs. quetiapine in patients prescribed simvastatin or atorvastatin

The proportional hazard assumption held for all four 12-month outcomes (Supplementary Figure 4). We found no evidence of an interaction between SMI diagnosis and trial arm for 12-month accident/injury admissions or 12-month self-harm events. There was evidence of an interaction between SMI diagnosis and trial arm for 12-month psychiatric (Wald test: p=0.002) and physical health (Wald test: p=0.010) admissions.

### Primary outcome: Psychiatric hospital admissions at 12 months

In patients prescribed simvastatin or atorvastatin, there was no difference in psychiatric admissions at 12 months between those initiating risperidone, olanzapine or aripiprazole compared to those initiating quetiapine (unadjusted HR: 0.97; 95%CI:0.83-1.12; adjusted HR: 0.93; 95%CI:0.79-1.09, Table 2, Figure 3). When stratified by SMI diagnosis there was no difference in psychiatric admissions between those initiating risperidone, olanzapine or aripiprazole compared to those initiating quetiapine for patients yet to receive a diagnosis (HR: 1.01; 95%CI:0.66-1.55) or with a diagnosis of bipolar disorder (HR:1.12; 95%CL:0.73-1.70). For those with a diagnosis of schizophrenia, we found a large negative effect size, though confidence intervals included the null (HR: 0.60; 95%CI:0.33-1.09). However, patients with other psychoses prescribed simvastatin or atorvastatin and initiating risperidone, olanzapine or aripiprazole had a lower rate of psychiatric admissions than those initiating quetiapine (HR: 0.53; 95%CI:0.34-0.85, Figure 4).

**Figure 3:**
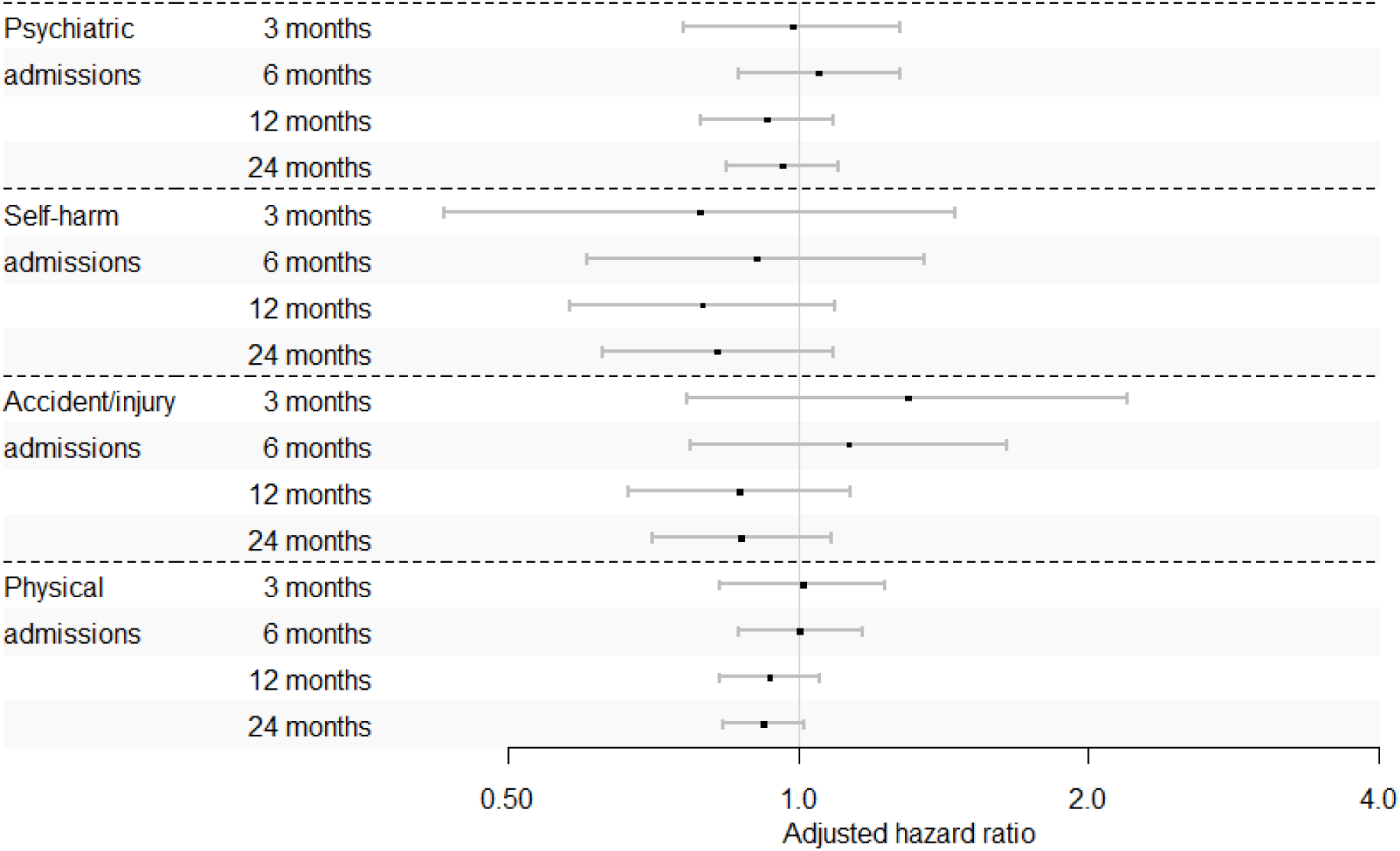
Cox regression results for trial 2B: Risperidone, olanzapine and aripiprazole vs. quetiapine in patients prescribed simvastatin or atorvastatin.

**Figure 4:**
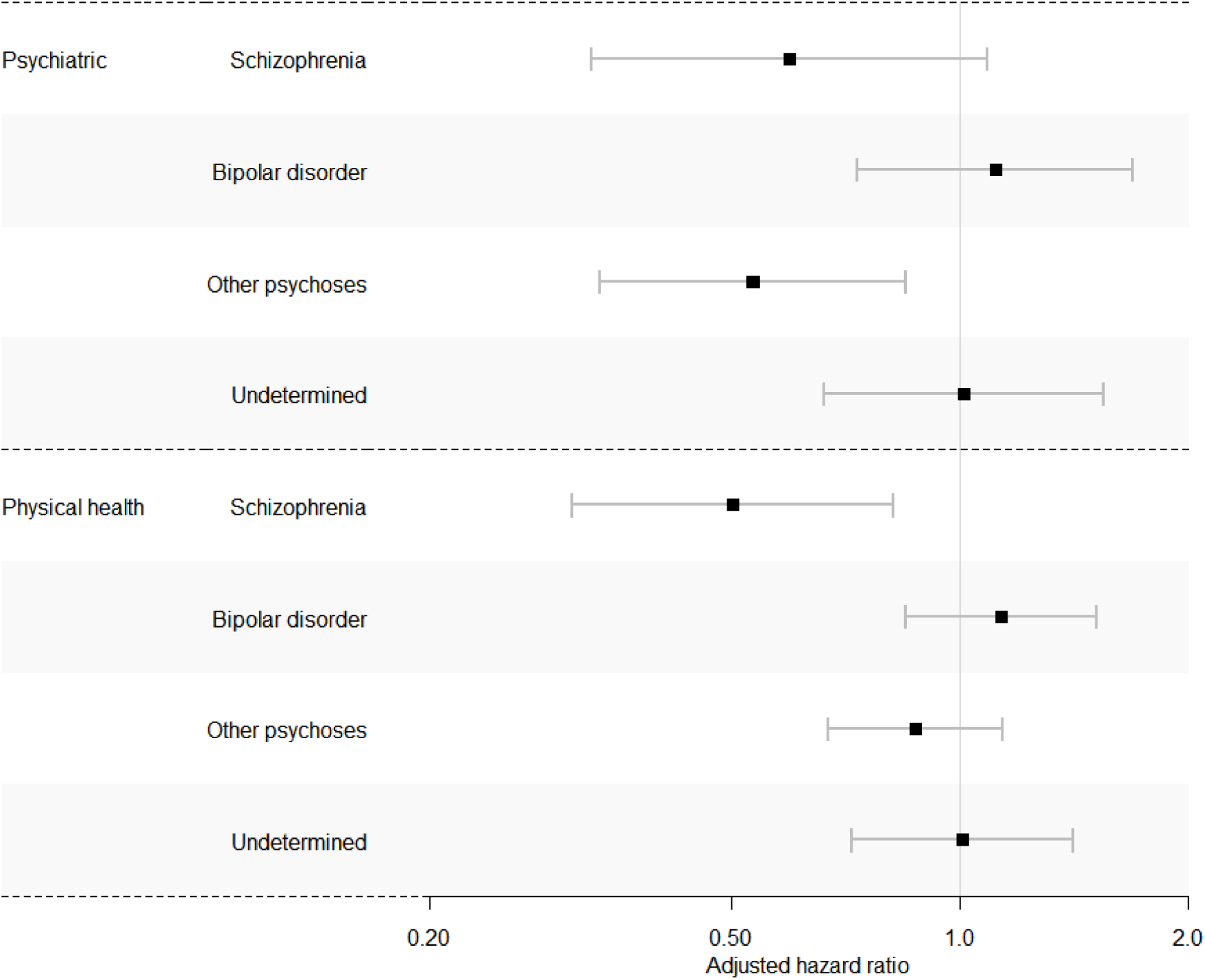
Stratified Cox regression results for Trial 2B: 12-month outcomes stratified by SMI diagnosis.

### Secondary outcomes

We observed no differences in psychiatric admissions at 3, 6 or 24 months and no difference in physical health or accident/injury admissions at any time point. Self-harm events were less frequent in people initiating risperidone, olanzapine or aripiprazole compared to quetiapine at 12 and 24 months in unadjusted analysis but not in adjusted analysis (Table 2, Figure 3). In stratified analysis, there was no difference in 12-month physical health admissions between trial arms for patients yet to receive a diagnosis (HR: 1.01; 95%CI:0.72-1.41), with a diagnosis of bipolar disorder (HR:1.14; 95%CL:0.85-1.52) or of other psychoses (HR: 0.87; 95%CI:0.0.67-1.14). However, patients with schizophrenia prescribed simvastatin or atorvastatin and initiating risperidone, olanzapine or aripiprazole had a lower rate of physical health admissions than those initiating quetiapine (HR: 0.50; 95%CI:0.31-0.82, Figure 4).

### Negative binomial models and sensitivity analyses

The adjusted analysis using negative binomial models found lower physical health admissions at 24 months in those initiating risperidone, olanzapine or aripiprazole compared to quetiapine (IRR: 0.90; 95%CI:0.81-0.99) and evidence of a lower rate of psychiatric admissions at 24 months, though the confidence interval was close to the null (IRR:0.87; 95%CI:0.76-0.999; Supplementary table 2). In contrast, the IPW model found no significant differences across any of the outcomes (Supplementary table 3). In per-protocol analysis we found evidence of a lower rate of self-harm events at 12 months in those initiating risperidone, olanzapine or aripiprazole compared to quetiapine (HR:0.60; 95%CI:0.38-0.97; supplementary table 4).

## Discussion

### Main findings

We conducted three TTE studies to test different hypotheses about the psychiatric effects of statins in patients with SMI: 1) BBB permeable statins vs non-permeable statins, in patients prescribed any mood stabiliser or antipsychotic 2A) statins inhibiting p-glycoprotein vs statins that do not in patients co-prescribed antipsychotics that are p-glycoprotein substrates, 2B) antipsychotics that are p-glycoprotein substrates vs antipsychotics that are not, in people co-prescribed statins that inhibit p-glycoprotein. We did not find between group differences in our primary endpoint; psychiatric admission by 12 months, in any of these comparisons.

We found no evidence to support the hypothesis that simvastatin may reduce psychiatric admissions in patients with SMI, compared to statins with less potential to cross the BBB. In fact, we found slightly elevated rates of psychiatric admission in patients prescribed simvastatin before confounder adjustment. Our secondary outcomes, and binomial and per protocol sensitivity analyses suggested a weak signal that patients prescribed simvastatin had increased rates of psychiatric admission, self-harm and accident/injury admissions compared to those prescribed atorvastatin, pravastatin or rosuvastatin.

We found no evidence to support the hypothesis that simvastatin and atorvastatin may reduce psychiatric admissions compared to pravastatin, in patients prescribed risperidone, aripiprazole or olanzapine. However, this study was statistically underpowered due to the low prevalence of pravastatin prescribing.

We found some evidence to support the hypothesis that patients prescribed simvastatin and atorvastatin may have reduced rates of psychiatric admission when prescribed risperidone, olanzapine or aripiprazole rather than quetiapine. However, this was only in the stratified analysis of the primary end point, and relating to the non-organic non-affective psychoses subgroup. The effect size was similar in the schizophrenia group (HR:0.53 in schizophrenia, and 0.6 in other psychoses), but confidence intervals were wide and contained the null. Patients prescribed these statins in combination with risperidone, olanzapine or aripiprazole also had lower rates of self-harm at 12 months compared to those prescribed quetiapine in the per-protocol analysis.

Taken together, these studies suggest that the psychiatric mechanism of action of statins is unlikely to be primarily via BBB permeability. P-glycoprotein inhibition increasing the effects of certain antipsychotics is a more probable mechanism of action, though we only identified effects in our primary end-point for patients with a diagnosis of ‘other psychosis’. Further research is therefore required to clarify the role of p-glycoprotein

### Comparison with existing literature and potential mechanisms

Our study found no improvement in psychiatric admissions in people prescribed simvastatin versus other statins. This is despite simvastatin having the most potential to cross the BBB based on the combination of molecular weight and lipophilicity and the most permeability *in vitro* (Sierra et al., 2011). Most RCTs of adjunct statins have focused on simvastatin, though findings have been mixed (Sommer et al., 2021; Tajik-Esmaeeli et al., 2017; Chaudhry et al., 2014; Aichholzer et al., 2022; Vincenzi et al., 2014; Ghanizadeh et al., 2014; Sayyah et al., 2015; Weiser et al., 2023). Our result concurs with those of Postolache et al., who found reduced psychiatric admissions in patients prescribed hydrophilic rather than lipophilic statins (Postolache et al., 2021), though our study defined BBB permeability beyond lipophilicity. Postolache et al., postulated that their findings could be *due* to their ability to cross the BBB, thereby lowering cholesterol in the brain, affecting serotonin synthesis, and increasing cognitive dysfunction, impulsivity and anxiety (Postolache et al., 2021). Another theory is that adjunct simvastatin is only effective in some patients (Zaki et al., 2024; Aichholzer et al., 2022).

Our results suggest that p-glycoprotein inhibition is a possible mechanism by which statins could improve psychiatric symptoms. If this is the case, we would expect to see improvement in psychiatric symptoms only when statins which inhibit p-glycoprotein (simvastatin and atorvastatin) are prescribed in combination with antipsychotics with an affinity for p-glycoprotein (such as aripiprazole, risperidone and olanzapine). This may explain some of the mixed results in published simvastatin trials. For example, two RCTs included patients on any antipsychotic agent, with simvastatin as an adjunct, and similar to our first TTE found no change in admissions at 12 months (Sommer et al., 2021; Weiser et al., 2023). In contrast, published RCTs which were limited to patients prescribed risperidone with or without simvastatin and risperidone with or without atorvastatin did find improvements in negative symptoms (Sayyah et al., 2015; Tajik-Esmaeeli et al., 2017). However, these trials were short term and of limited sample size. The results of these trials plus our observational TTEs suggest that larger trials are warranted to investigate whether simvastatin or atorvastatin in combination with aripiprazole, risperidone and olanzapine may improve psychiatric symptoms.

However, an RCT of pravastatin, which does not readily cross the BBB and does not have affinity for p-glycoprotein also found a small reduction in short-term positive symptoms at 6 weeks in people with schizophrenia (Vincenzi et al., 2014). If this effect is true for pravastatin, it suggests that mechanisms other than those tested in our studies may also play a role in the improvement of psychiatric symptoms following statin initiation. There is therefore a need for more research into the mechanisms of action and potential interactions between individual statins and antipsychotics.

### Strengths and Limitations

For the first time we used large, representative electronic health records and TTE to estimate the causal effect of statin/psychotropic combinations on psychiatric admissions in patients with SMI. We used this approach to test three mechanistic hypotheses.

The use of a TTE framework is a major advantage of this study. By pre-registering our protocol, designing a hypothetical RCT and emulating that using observational data we sought to reduce potential biases introduced by observational study design (Hernan et al., 2022). There are likely to be large differences between those who are and aren’t prescribed statins, many of which will not be measurable using electronic health records and therefore near impossible to account for. An advantage of the head-to-head design of our study is the reduction in confounding by indication compared to observational study designs comparing those with and without prescriptions for statins, as indication for statins is required for all arms of the analysis. While previous self-controlled study analyses remove much of this, time varying confounding, such as cardiovascular events may still confound the relationship between statin prescription and psychiatric outcome. Observational studies in the general population have found a higher rate of suicide and depression following the initiation of statins (Ye et al., 2023), likely due to cardiovascular events that precipitate their prescription. Alternatively, those prescribed statins may be more likely to access healthcare, and may have better adherence to antipsychotics (MacEwan et al., 2018), thereby reducing the risk of psychiatric outcomes. The head-to-head TTEs we describe overcome these problems with previous observational studies.

Despite the large size of our original cohort, Trial 2A in particular was underpowered. This may explain the negative finding. For Trial 1 and 2B, the sample size was greater than most RCTs on the subject. However, while we determined the cohort would be large enough to detect a 10% change in hospitalisation rates (see protocol), and previous studies have found a change in admissions of between 20 and 30% (Hayes et al., 2019; Postolache et al., 2021), the imbalance in sample size between treatment arms and the over-dispersed nature of hospital admissions means that our null finding could be due to issues of power. Furthermore, our outcomes were limited to outcomes that are readily available in electronic health records. We therefore studied psychiatric admissions and self-harm events as proxies of psychiatric functioning. These outcomes are not as sensitive as symptom severity, and we therefore miss patients who show worsening of symptoms which do not result in either hospitalisation or self-harm events.

Some unmeasured confounding may have remained in our studies. For example, those prescribed high-intensity statins may have worse cardiovascular health and therefore be more likely to experience the psychiatric outcomes. While we adjusted for statin intensity, some residual confounding by indication may remain. Furthermore, our work was based on primary care records and therefore, while we had data on prescriptions issued to patients, we did not have data on whether these medications were dispensed or taken. Our measures of prescriptions therefore likely over-estimates the number of patients who adhere to medication. However, since all patients had to be on both antipsychotics and statins, the differences are unlikely to be differential between our active and comparator arms. There are a range of patient, provider and societal factors that influence the recording of self-harm events, and the frequency of psychiatric admissions. While we were not able to measure all of these factors, it is unlikely that these factors were differential across active and comparator arms.

We found some weak evidence of reduced self-harm events and psychiatric admissions following initiation of risperidone, olanzapine or aripiprazole in people prescribed simvastatin and atorvastatin. While a recent study found no differences in psychiatric admissions between patients prescribed these four antipsychotics (Richards-Belle et al., 2024), we cannot rule out the possibility that these results are due to differences in the action of these antipsychotics, rather than the combination of antipsychotic plus statin.

While we formed our hypotheses based on previous work, it is possible that certain statin-antipsychotic combinations may be effective through mechanisms not considered in this study. We were unable to investigate the effects of individual statin-antipsychotic combinations, which may have resulted in a dilution of the true effects. Furthermore, our strict inclusion and exclusion criteria limited the size and generalizability of the populations we included. Notably, our study cohort were far older than other cohorts of patients with SMI, and our analysis was under-powered to investigate some combinations of statins and antipsychotic medications. Also, our study required patients to be eligible for linkage to both CPRD and HES. While the majority of patients in CPRD are eligible for linkage, and CPRD has been shown to be broadly representative of the population of England(Wolf et al., 2019; Herrett et al., 2015), biases introduced by this selection process cannot be ruled out.

Our study did not compare the overall effects of statin prescription on psychiatric outcomes, but sought to identify differences between statins to identify targets for future research and convey information on hypothesised modes of action. Comparison to placebo is best approached through interventional study designs.

### Conclusions

We found no evidence to support the use of simvastatin as an adjunct to antipsychotics in people with severe mental illness. However, we found weak evidence that for those prescribed risperidone, olanzapine or aripiprazole, the addition of simvastatin or atorvastatin may reduce psychiatric admissions and self-harm. The results of this study suggest a need to conduct RCTs that focus on individual statin and antipsychotic combinations, and in particular those statins and antipsychotics which interact with p-glycoprotein; as well as further studies into the mechanisms of action and interaction of statins and antipsychotics.

## Statements and declarations

### Ethical considerations

Approval for this study was obtained from the Independent Scientific Advisory Committee of CPRD (protocol number 21_000729). CPRD obtains annual research ethics approval from the UK Health Research Authority Research Ethics Committee (East Midlands—Derby Research Ethics Committee reference number 05/MRE04/87) to receive and supply patient data for public health research. Therefore, no additional ethics approval was required for this study.

### Consent to participate

CPRD uses anonymised patient records and therefore does not require individual patient consent. However, patients are able to opt out of their medical records being used for research.

### Declaration of competing interests

NL & ARB declare no conflicts of interest.

JFH has received consultancy fees from juli Health and Wellcome. He is a co-founder and shareholder of juli Health. juli Health has a patent pending.

ICKW reports research funding from Amgen, Bristol Myers Squibb, Pfizer, Janssen, Bayer, GSK, Novartis, the Hong Kong Research Grants Council of the Government of the Hong Kong SAR, the Hong Kong Health and Medical Research Fund, the National Institute for Health Research in England, the European Commission, and the National Health and Medical Research Council in Australia, consulting fees from IQVIA and World Health Organisation, payment for expert testimony for Appeal Court of Hong Kong and is a non-executive director of Jacobson Medical in Hong Kong, Advance Data Analytics for Medical Science (ADAMS) Limited (HK), Asia Medicine Regulatory Affairs (AMERA)

Services Limited and OCUS Innovation Limited (HK, Ireland and UK) a former director of Therakind in England, outside of the submitted work.

KKCM reports personal fees from IQVIA, unrelated to the current work.

### Funding statement

This work is supported by the UK Research and Innovation grant MR/V023373/1 (NL&JFH). It is additionally supported by UK Research and Innovation grant MR/W014386/1 (NL&DPJO), the University College London Hospitals NIHR Biomedical Research Centre (NL, JFH & DPJO), and the NIHR North Thames Applied Research Collaboration (NL, JFH & DPJO). These funders had no role in study design, data collection, data analysis, data interpretation, or writing of the report. The views expressed in this article are those of the authors and not necessarily those of the NHS, the NIHR, or the Department of Health and Social Care.

NL is supported by an Health Data Research UK personal fellowship., This work is affiliated to Health Data Research UK (Big Data for Complex Disease-HDR-23012), which is funded by the Medical Research Council (UKRI), the National Institute for Health Research, the British Heart Foundation, Cancer Research UK, the Economic and Social Research Council (UKRI), the Engineering and Physical Sciences Research Council (UKRI), Health and Care Research Wales, Chief Scientist Office of the Scottish Government Health and Social Care Directorates, and Health and Social Care Research and Development Division (Public Health Agency, Northern Ireland).

ICKW is partially supported by the Laboratory of Data Discovery for Health (D24H) funded by the by AIR@InnoHK administered by Innovation and Technology Commission, Hong Kong and received research funding from Amgen, Bristol Myers Squibb, Pfizer, Janssen, Bayer, GSK, Novartis, the Hong Kong Research Grants Council of the Government of the Hong Kong SAR, the Hong Kong Health and Medical Research Fund, the National Institute for Health Research in England, the European Commission, and the National Health and Medical Research Council in Australia, consulting fees from IQVIA and World Health Organisation, payment for expert testimony for Appeal Court of Hong Kong and is a non-executive director of Jacobson Medical in Hong Kong, Advance Data Analytics for Medical Science (ADAMS) Limited (HK), and OCUS Innovation Limited (HK, Ireland and UK) a former director of Therakind in England, outside of the submitted work.

ARB is funded by the Wellcome Trust through a PhD Fellowship in Mental Health Science (218497/Z/19/Z). This research was funded in whole or in part by the Wellcome Trust. For the purpose of Open Access, the author has applied a CC BY public copyright licence to any Author Accepted Manuscript (AAM) version arising from this submission.

KKCM reports grants from the CW Maplethorpe Fellowship, the European Union Horizon 2020, the UK National Institute of Health Research, the Hong Kong Research Grant Council, and the Hong Kong Innovation and Technology Commission.

### Data Availability

Electronic health records are, by definition, considered to be sensitive data in the UK by the Data Protection Act and cannot be shared via public deposition because of information governance restriction in place to protect patient confidentiality. Access to data is available only once approval has been obtained through the individual constituent entities controlling access to the data. The primary care data can be requested via application to the Clinical Practice Research Datalink.

## Notes

### Clinical Protocols

https://osf.io/hck8n

### Author Declarations

Approval for this study was obtained from the Independent Scientific Advisory Committee of CPRD (protocol number 21 000729). CPRD obtains annual research ethics approval from the UK Health Research Authority Research Ethics Committee (East Midlands Derby Research Ethics Committee reference number 05/MRE04/87) to receive and supply patient data for public health research. Therefore, no additional ethics approval was required for this study. CPRD uses anonymised patient records and therefore does not require individual patient consent. However, patients are able to opt out of their medical records being used for research.

