## Supplementary material for "Psychiatric symptom improvement from adjunctive statin prescribing in severe mental illness: three target trial emulation studies"

### Supplementary materials

##### Supplementary Table 1: Hypothetical ideal trial and observational analogues.

*Trial 1:* *Simvastatin vs. atorvastatin, pravastatin or rosuvastatin in people with SMI prescribed antipsychotics or mood stabilisers*

|  | **Target trial** | **Emulated trial** |
| --- | --- | --- |
| Inclusions | Adults (>18 years at study entry) with a diagnosis of SMI prior to entry and with at least 6 months of psychotropic medication (antipsychotic, lithium, sodium valproate or lamotrigine) use at a therapeutic dose with newly diagnosed dyslipidaemia for which a statin is indicated. | Adults (>18 years at study entry) with a diagnosis of SMI recorded in their EHR with a first prescription for statins and with at least one prescription for psychotropic medication (antipsychotic, lithium, sodium valproate or lamotrigine) at a therapeutic dose in the 6 months prior to statin prescription. |
| Exclusions | - Prior prescriptions of statins - Dyslipidaemia prior to study entry - Injectable antipsychotics in the 3 months prior to study entry - Contraindications to statins (hepatic impairment and myopathy) | - Prior prescriptions of statins - Injectable antipsychotics in the 3 months prior to study entry or antipsychotics not at therapeutic dose^a^. - Contraindications to statins can be assumed to be absent given they will be prescribed statins in clinical practice. |
| Treatment strategies | 1) Initiate simvastatin 2) Initiate atorvastatin, pravastatin or rosuvastatin | Same |
| Assignment procedure | Randomly assigned at baseline to treatment or comparison arms, but aware of their status. | Assigned at study entry to treatment or comparison arms based on the initiation of statins as recorded in their EHR, with randomisation emulated through controlling of confounders. |
| Recruitment period | Jan 2000 to December 2018. | Same. |
| Start of follow up | Randomisation which must occur within three months of diagnosis of dyslipidaemia. | First prescription of statin in primary care |
| End of follow up | Outcome of interest (for time-to-event), loss to follow up, two years of follow up, or December 2019. | Same. |
| Outcome | 1) Psychiatric admission 2) Self-harm events 3) Physical health admission 4) Accident or injury admission | Same. |
| Causal contrast of interest | Intention to treat effect. | Observational analogue of intention to treat effect. |
| Analyses | Hazard ratio and incidence rate ratio | Same. |

*Trial 2A: Simvastatin or atorvastatin vs. pravastatin in people with SMI prescribed risperidone, aripiprazole or olanzapine*

|  | **Target trial** | **Emulated trial** |
| --- | --- | --- |
| Inclusions | Adults (>18 years at study entry) with a diagnosis of SMI prior to entry, and with at least 6 months of risperidone, aripiprazole or olanzapine use at a therapeutic dose, and newly diagnosed dyslipidaemia for which a statin is indicated. | Adults (>18 years at study entry) with a diagnosis of SMI recorded in their EHR with a first prescription for simvastatin, atorvastatin or pravastatin and with at least one prescription for risperidone, olanzapine or aripiprazole at a therapeutic dose in the 6 months prior to statin prescription. |
| Exclusions | As per target trial 1. | As per emulated trial 1. |
| Treatment strategies | 1) Initiate simvastatin 2) Initiate atorvastatin  3) Initiate pravastatin | Same. |
| Assignment procedure | As per target trial 1. | As per emulated trial 1. |
| Recruitment period | As per target trial 1. | As per emulated trial 1. |
| Start of follow up | As per target trial 1. | As per emulated trial 1. |
| End of follow up | As per target trial 1. | As per emulated trial 1. |
| Outcome | As per target trial 1. | As per emulated trial 1. |
| Causal contrast of interest | As per target trial 1. | As per emulated trial 1. |
| Analyses | As per target trial 1. | As per emulated trial 1. |

*Trial 2B: Risperidone, olanzapine and aripiprazole vs. quetiapine in patients prescribed simvastatin or atorvastatin*

|  | **Target trial** | **Emulated trial** |
| --- | --- | --- |
| Inclusions | Adults (>18 years at study entry) with a diagnosis of dyslipidaemia prior to entry and a diagnosis SMI for which risperidone, aripiprazole, olanzapine or quetiapine is indicated. | Adults (>18 years at study entry) newly prescribed risperidone, aripiprazole, olanzapine or quetiapine with a statin prescription recorded in their EHR in the six months before the newly prescribed antipsychotic. |
| Exclusions | - Prior prescriptions of risperidone, aripiprazole, olanzapine or quetiapine. - Injectable antipsychotics in the 3 months prior to study entry. - Contraindications to olanzapine (acute myocardial infarction, bradycardia, recent heart surgery, severe hypotension, sick sinus syndrome or unstable angina) | - Prior prescriptions of risperidone, aripiprazole, olanzapine or quetiapine. - Injectable antipsychotics in the 3 months prior to study entry. - Contraindications to risperidone, aripiprazole, olanzapine or quetiapine   can be assumed to be absent given they will be prescribed these in clinical practice. |
| Treatment strategies | 1) Initiate risperidone, aripiprazole or olanzapine. 2) Initiate quetiapine. | Same. |
| Assignment procedure | As per target trial 1. | As per emulated trial 1. |
| Recruitment period | As per target trial 1. | As per emulated trial 1. |
| Start of follow up | Initiation of risperidone, aripiprazole, olanzapine or quetiapine | One month after the initiation of risperidone, aripiprazole, olanzapine or quetiapine to allow titration to therapeutic dose |
| End of follow up | As per target trial 1 | As per emulated trial 1 |
| Outcome | As per target trial 1 | As per emulated trial 1 |
| Causal contrast of interest | As per target trial 1 | As per emulated trial 1 |
| Analyses | As per target trial 1 | As per emulated trial 1 |

##### Supplementary Table 2: Negative binomial results

|  | | | 3 months,  hazard ratio (95% CI) | 6 months,  hazard ratio (95% CI) | 12 months, hazard ratio (95% CI) | 24 months,  hazard ratio (95% CI) |
| --- | --- | --- | --- | --- | --- | --- |
| TRIAL 1: Simvastatin vs. atorvastatin, pravastatin or rosuvastatin in people with SMI prescribed antipsychotics or mood stabilisers | | | | | | |
|  | Psychiatric admissions | Unadjusted | 1.20 (0.94-1.53) | 1.12 (0.94-1.33) | 1.10 (0.96-1.27) | 1.07 (0.95-1.20) |
|  |  | Adjusted | 1.04 (0.71-1.53) | 0.92 (0.69-1.22) | 0.98 (0.78-1.25) | 0.89 (0.74-1.08) |
|  | Self-harm events | Unadjusted | 0.86 (0.54-1.37) | 0.83 (0.59-1.16) | 0.85 (0.65-1.11) | 0.87 (0.70-1.08) |
|  |  | Adjusted | 1.95 (1.03-3.70) | 1.32 (0.79-2.21) | 1.19 (0.79-1.80) | 1.03 (0.75-1.41) |
|  | Physical health admissions | Unadjusted | 0.87 (0.75-1.02) | 0.91 (0.81-1.03) | 0.92 (0.84-1.02) | 0.92 (0.84-0.99) |
|  |  | Adjusted | 1.06 (0.84-1.32) | 1.04 (0.86-1.24) | 1.09 (0.94-1.26) | 1.00 (0.88-1.14) |
|  | Accident/injury admissions | Unadjusted | 1.02 (0.65-1.59) | 1.07 (0.80-1.44) | 1.02 (0.83-1.26) | 1.07 (0.91-1.25) |
|  |  | Adjusted | 1.26 (0.57-2.77) | 1.41 (0.86-2.29) | 1.09 (0.77-1.56) | 1.12 (0.87-1.43) |
| TRIAL 2A: Simvastatin or atorvastatin vs. pravastatin in people with SMI prescribed risperidone, aripiprazole or olanzapine | | | | | | |
|  | Psychiatric admissions | Unadjusted | 0.73 (0.17- 3.21) | 0.54 (0.21-1.38) | 0.58 (0.26-1.32) | 0.74 (0.36-1.51) |
|  |  | Adjusted | 0.53 (0.11-2.56) | 0.52 (0.17-1.60) | 0.65 (0.27-1.60) | 0.89 (0.42-1.86) |
|  | Self-harm events | Unadjusted |  | 2.42 (0.17-34.20) | 5.17 (0.45-58.96) | 4.99 (0.79-31.60) |
|  |  | Adjusted |  | 1.79 (0.23-14.23) | 6.11 (0.88-42.34) | 4.24 (0.85-21.22) |
|  | Physical health admissions | Unadjusted | 1.43 (0.40- 5.19) | 1.26 (0.49-3.21) | 1.07 (0.52-2.18) | 1.01 (0.57-1.82) |
|  |  | Adjusted | 1.89 (0.58-6.16) | 1.47 (0.63-3.40) | 1.27 (0.65-2.50) | 1.17 (0.68-2.00) |
|  | Accident/injury admissions | Unadjusted |  |  | 2.09 (0.26-16.89) | 2.00 (0.45-8.97) |
|  |  | Adjusted |  |  | 2.32 (0.32-16.83) | 1.81 (0.44-7.41) |
| TRIAL 2B: Risperidone, olanzapine and aripiprazole vs. quetiapine in patients prescribed simvastatin or atorvastatin | | | | | | |
|  | Psychiatric admissions | Unadjusted | 0.90 (0.72-1.11) | 1.00 (0.84-1.19) | 0.89 (0.77-1.03) | 0.91 (0.80-1.04) |
|  |  | Adjusted | 0.86 (0.68-1.09) | 0.99 (0.82-1.19) | 0.85 (0.73-1.00) | 0.87 (0.76-0.999) |
|  | Self-harm events | Unadjusted | 0.60 (0.37-0.99) | 0.70 (0.47-1.04) | 0.77 (0.55-1.08) | 0.80 (0.59-1.08) |
|  |  | Adjusted | 0.68 (0.41-1.14) | 0.95 (0.64-1.43) | 0.96 (0.69-1.33) | 1.04 (0.77-1.41) |
|  | Physical health admissions | Unadjusted | 1.01 (0.85-1.21) | 1.02 (0.89-1.17) | 0.99 (0.88-1.11) | 0.98 (0.88-1.08) |
|  |  | Adjusted | 0.92 (0.77-1.10) | 0.95 (0.82-1.09) | 0.90 (0.79-1.01) | 0.90 (0.81-0.99) |
|  | Accident/injury admissions | Unadjusted | 0.95 (0.64-1.41) | 1.09 (0.79-1.49) | 0.86 (0.69-1.08) | 0.84 (0.70-1.02) |
|  |  | Adjusted | 0.97 (0.65-1.47) | 1.08 (0.78-1.50) | 0.86 (0.68-1.07) | 0.85 (0.71-1.01) |

##### Supplementary Table 3: Inverse probability weighted models

|  | | | 3 months,  hazard ratio (95% CI) | 6 months,  hazard ratio (95% CI) | 12 months,  hazard ratio (95% CI) | 24 months,  hazard ratio (95% CI) |
| --- | --- | --- | --- | --- | --- | --- |
| TRIAL 1: Simvastatin vs. atorvastatin, pravastatin or rosuvastatin in people with SMI prescribed antipsychotics or mood stabilisers | | | | | | |
|  | Psychiatric admissions | Weighted | 1.49 (1.13-1.98) | 1.13 (0.93-1.36) | 1.13 (0.98-1.30) | 1.06 (0.95-1.19) |
|  |  | Weighted & adjusted^1^ | 1.46 (1.08-1.97) | 1.06 (0.87-1.30) | 1.11 (0.95-1.31) | 1.02 (0.90-1.16) |
|  | Self-harm events | Weighted | 1.56 (1.00-2.44) | 0.94 (0.60-1.45) | 0.94 (0.70-1.27) | 0.88 (0.69-1.11) |
|  |  | Weighted & adjusted^1^ | 1.70 (1.05-2.76) | 0.89 (0.56-1.42) | 0.91 (0.64-1.30) | 0.81 (0.62-1.05) |
|  | Physical health admissions | Weighted | 0.97 (0.81-1.16) | 0.98 (0.84-1.15) | 0.99 (0.88-1.13) | 0.97 (0.87-1.06) |
|  |  | Weighted & adjusted^1^ | 1.02 (0.82-1.27) | 1.04 (0.86-1.25) | 1.04 (0.89-1.21) | 0.98 (0.87-1.10) |
|  | Accident/injury admissions | Weighted | 1.56 (0.85-2.85) | 1.45 (1.01-2.09) | 1.10 (0.81-1.50) | 1.09 (0.86-1.37) |
|  |  | Weighted & adjusted^1^ | 1.75 (0.89-3.44) | 1.62 (1.05-2.50) | 1.14 (0.80-1.63) | 1.10 (0.85-1.44) |
| TRIAL 2A: Simvastatin or atorvastatin vs. pravastatin in people with SMI prescribed risperidone, aripiprazole or olanzapine | | | | | | |
|  | Psychiatric admissions | | 5.88 (0.15-232.56) | 3.77 (0.14-100.55) | 1.75 (0.10-31.00) | 1.89 (0.22-16.10) |
|  | Self-harm events | |  | 0.94 (0.05-16.34) | 1.70 (0.15-19.07) | 0.80 (0.14-4.47) |
|  | Physical health admissions | | 19.19 (1.03-357.33) | 21.20 (1.71-262.51) | 12.04 (1.60-90.80) | 0.59 (0.27-1.31) |
|  | Accident/injury admissions | |  |  | 3.26 (0.30-35.82) | 0.57 (0.14-2.42) |
| TRIAL 2B: Risperidone, olanzapine and aripiprazole vs. quetiapine in patients prescribed simvastatin or atorvastatin | | | | | | |
|  | Psychiatric admissions | | 0.97 (0.77-1.21) | 1.01 (0.85-1.20) | 0.88 (0.76-1.00) | 0.91 (0.81-1.02) |
|  | Self-harm events | | 0.89 (0.59-1.34) | 0.91 (0.67-1.23) | 0.82 (0.64-1.04) | 0.82 (0.66-1.01) |
|  | Physical health admissions | | 0.99 (0.84-1.17) | 0.98 (0.86-1.11) | 0.93 (0.84-1.03) | 0.92 (0.84-1.00) |
|  | Accident/injury admissions | | 1.24 (0.80-1.91) | 1.20 (0.87-1.65) | 0.89 (0.71-1.13) | 0.92 (0.76-1.10) |

1: Additionally adjusted for statin dose

##### Supplementary Table 4: Per-protocol analysis

|  | | | 3 months,  hazard ratio (95% CI) | 6 months,  hazard ratio (95% CI) | 12 months,  hazard ratio (95% CI) | 24 months,  hazard ratio (95% CI) |
| --- | --- | --- | --- | --- | --- | --- |
| TRIAL 1: Simvastatin vs. atorvastatin, pravastatin or rosuvastatin in people with SMI prescribed antipsychotics or mood stabilisers | | | | | | |
|  | Psychiatric admissions | Unadjusted | 1.32 (1.02-1.70) | 1.23 (1.02-1.49) | 1.24 (1.06-1.44) | 1.16 (1.02-1.33) |
|  |  | Adjusted | 0.91 (0.60- 1.36) | 0.83 (0.62- 1.10) | 1.10 (0.86- 1.40) | 1.04 (0.85- 1.28) |
|  | Self-harm events | Unadjusted | 1.09 (0.73-1.61) | 0.94 (0.70-1.25) | 0.94 (0.74-1.18) | 0.93 (0.76-1.13) |
|  |  | Adjusted | 1.33 (0.60- 2.97) | 0.99 (0.57- 1.70) | 0.92 (0.61- 1.40) | 0.94 (0.66- 1.32) |
|  | Physical health admissions | Unadjusted | 0.93 (0.79-1.08) | 0.96 (0.85-1.09) | 0.99 (0.89-1.10) | 0.96 (0.88-1.05) |
|  |  | Adjusted | 1.11 (0.86- 1.43) | 1.05 (0.86- 1.28) | 1.12 (0.95- 1.32) | 1.08 (0.94- 1.24) |
|  | Accident/injury admissions | Unadjusted | 1.03 (0.65-1.64) | 1.12 (0.81-1.54) | 1.08 (0.84-1.38) | 1.07 (0.87-1.30) |
|  |  | Adjusted | 1.43 (0.56- 3.64) | 1.38 (0.77- 2.46) | 1.14 (0.74- 1.74) | 1.08 (0.78- 1.50) |
| TRIAL 2A: Simvastatin or atorvastatin vs. pravastatin in people with SMI prescribed risperidone, aripiprazole or olanzapine | | | | | | |
|  | Psychiatric admissions | Unadjusted | 0.94 (0.17-5.04) | 0.61 (0.18-2.10) | 0.61 (0.20-1.83) | 1.07 (0.38-3.03) |
|  |  | Adjusted | 0.36 (0.03-4.32) | 0.41 (0.08-2.01) | 0.41 (0.11-1.52) | 0.93 (0.27-3.24) |
|  | Self-harm events | Unadjusted |  | 0.52 (0.04-6.21) | 1.21 (0.13-11.42) | 1.68 (0.19-14.91) |
|  |  | Adjusted |  | 0.02 (0.00-25.02) | 1.23 (0.02-77.61) | 1.98 (0.08-52.18) |
|  | Physical health admissions | Unadjusted | 0.90 (0.24-3.41) | 0.80 (0.28-2.29) | 0.67 (0.28-1.57) | 0.85 (0.40-1.80) |
|  |  | Adjusted | 089 (0.17-4.69) | 0.55 (0.16-1.86) | 0.72 (0.26-2.02) | 0.89 (0.37-2.16) |
|  | Accident/injury admissions | Unadjusted |  |  |  | 0.79 (0.08-7.41) |
|  |  | Adjusted |  |  |  | 0.96 (0.04-22.31) |
| TRIAL 2B: Risperidone, olanzapine and aripiprazole vs. quetiapine in patients prescribed simvastatin or atorvastatin | | | | | | |
|  | Psychiatric admissions | Unadjusted | 1.02 (0.78-1.33) | 1.09 (0.87-1.38) | 0.94 (0.77-1.15) | 0.89 (0.73-1.07) |
|  |  | Adjusted | 0.97 (0.72- 1.32) | 1.06 (0.82-1.37) | 0.89 (0.72-1.12) | 0.85 (0.69-1.05) |
|  | Self-harm events | Unadjusted | 0.77 (0.46-1.26) | 0.74 (0.49-1.11) | 0.64 (0.44-0.91) | 0.62 (0.44-0.87) |
|  |  | Adjusted | 0.93 (0.43- 2.02) | 0.80 (0.46- 1.40) | 0.60 (0.38-0.97) | 0.67 (0.43-1.05) |
|  | Physical health admissions | Unadjusted | 1.08 (0.88-1.32) | 1.05 (0.88-1.24) | 1.07 (0.92-1.24) | 1.02 (0.89-1.17) |
|  |  | Adjusted | 0.97 (0.78- 1.22) | 0.97 (0.80-1.17) | 0.98 (0.83-1.15) | 0.92 (0.80-1.07) |
|  | Accident/injury admissions | Unadjusted | 1.26 (0.75-2.12) | 1.26 (0.82-1.95) | 1.01 (0.71-1.44) | 0.99 (0.72-1.35) |
|  |  | Adjusted | 1.25 (0.66- 2.36) | 1.34 (0.80-2.22) | 0.95 (0.63-1.43) | 0.91 (0.63-1.30) |

##### Supplementary Figure 1: Study inclusion flow chart

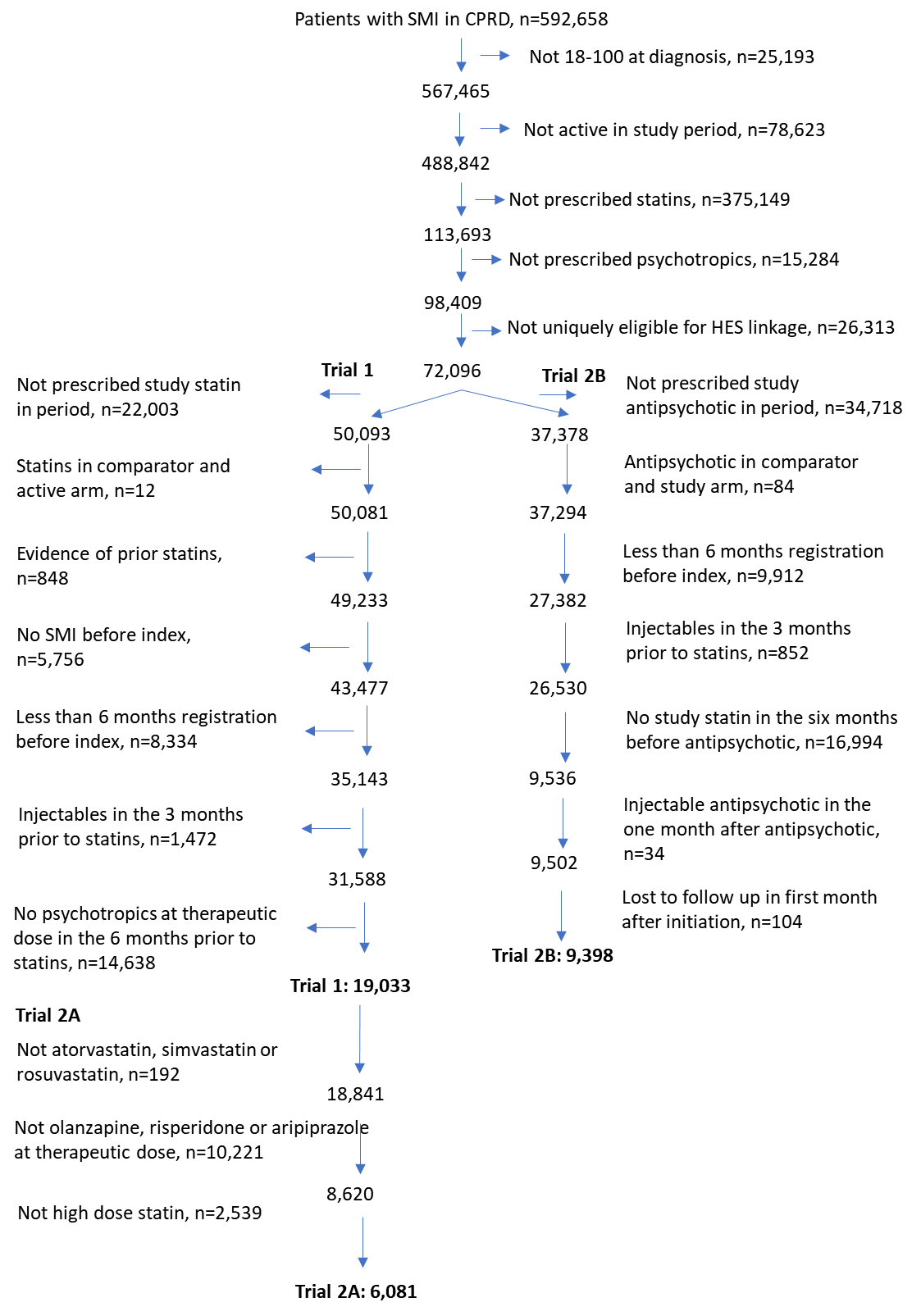

##### Supplementary Figure 2: Schoenfeld residual and Kaplan Meier plots, Trial 1

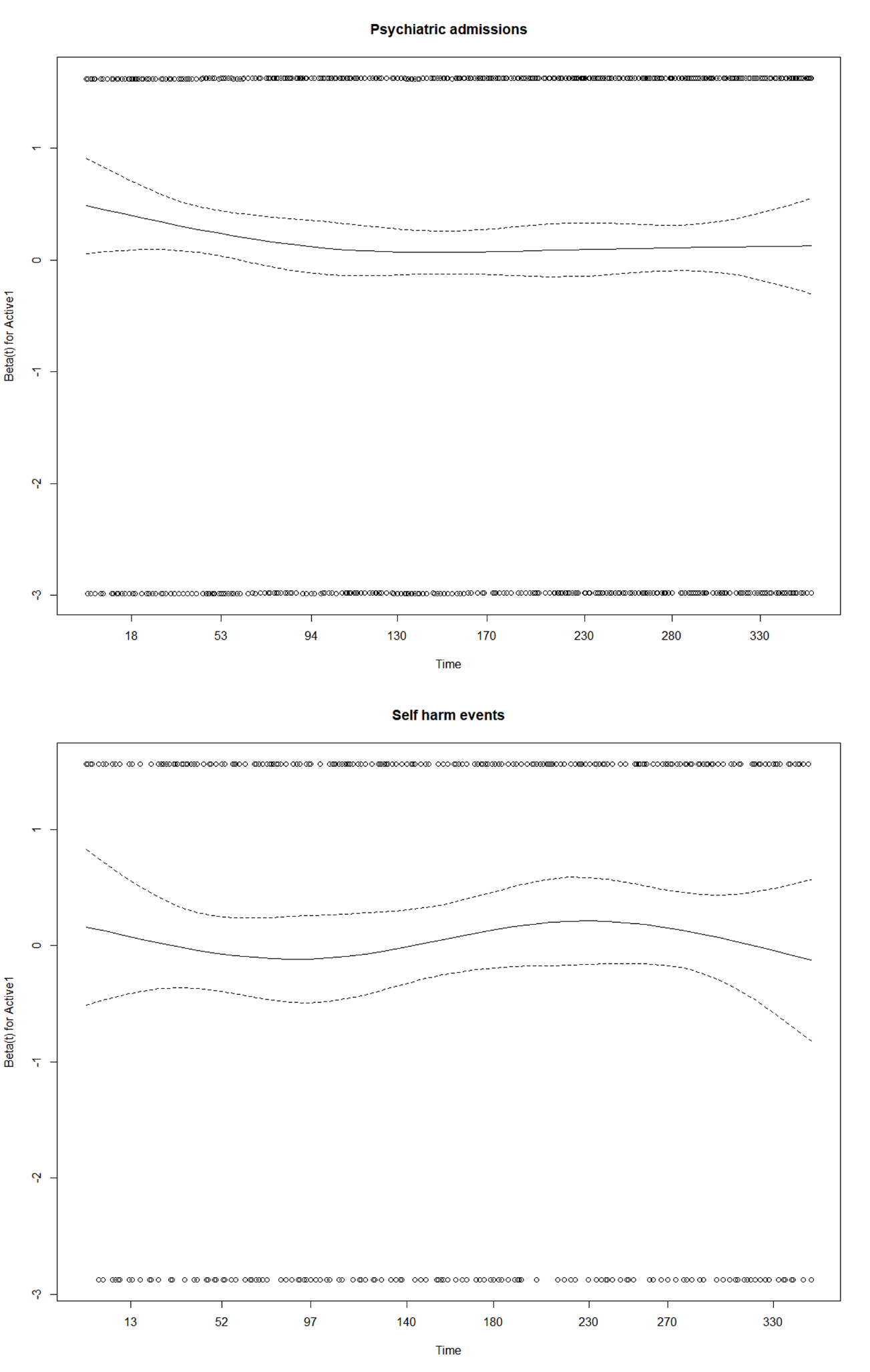

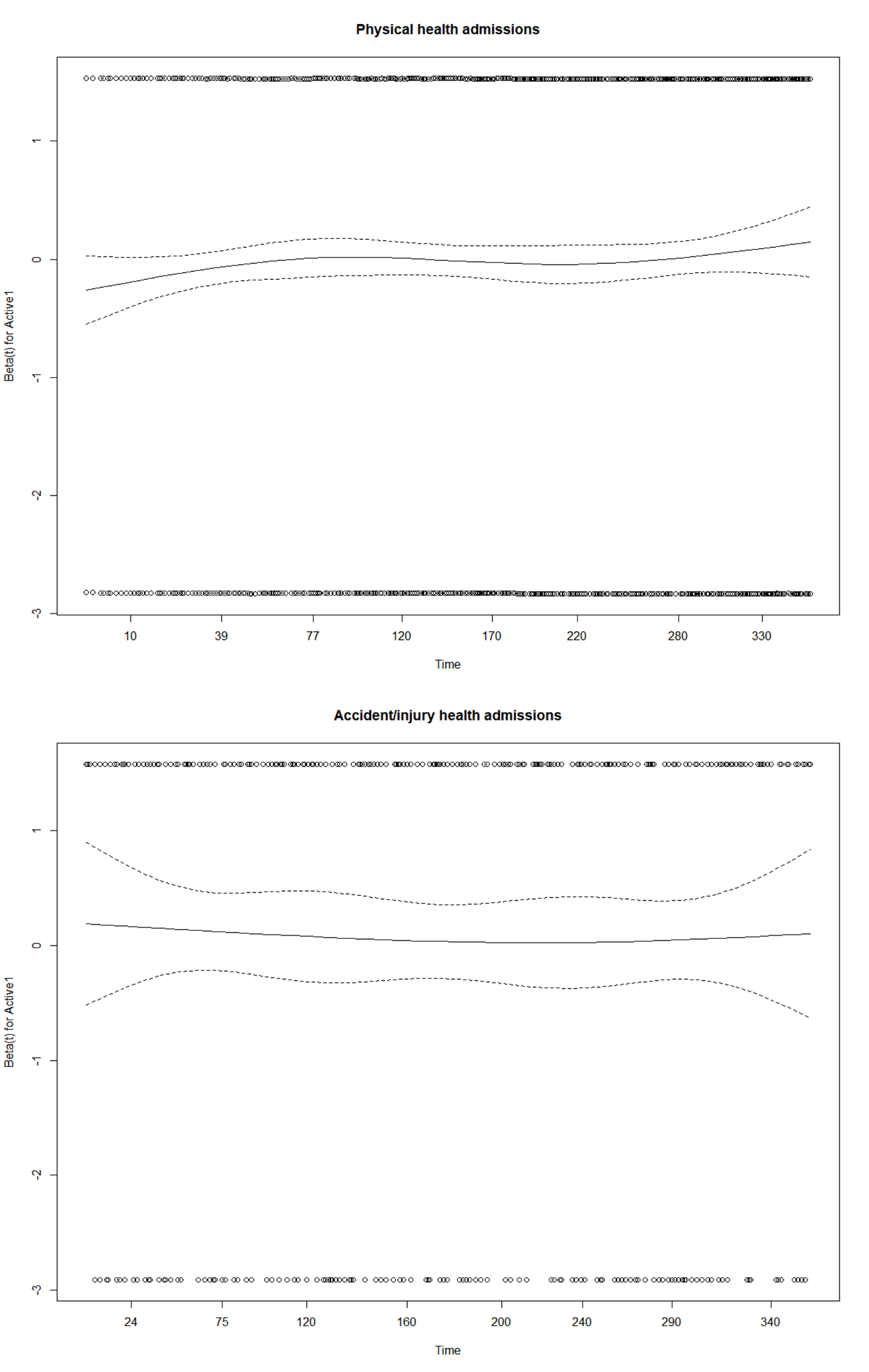

Psychiatric admissions: p=0.23; self-harm: p=0.71; physical health admissions: p=0.13; accident and injury admissions: p=0.76

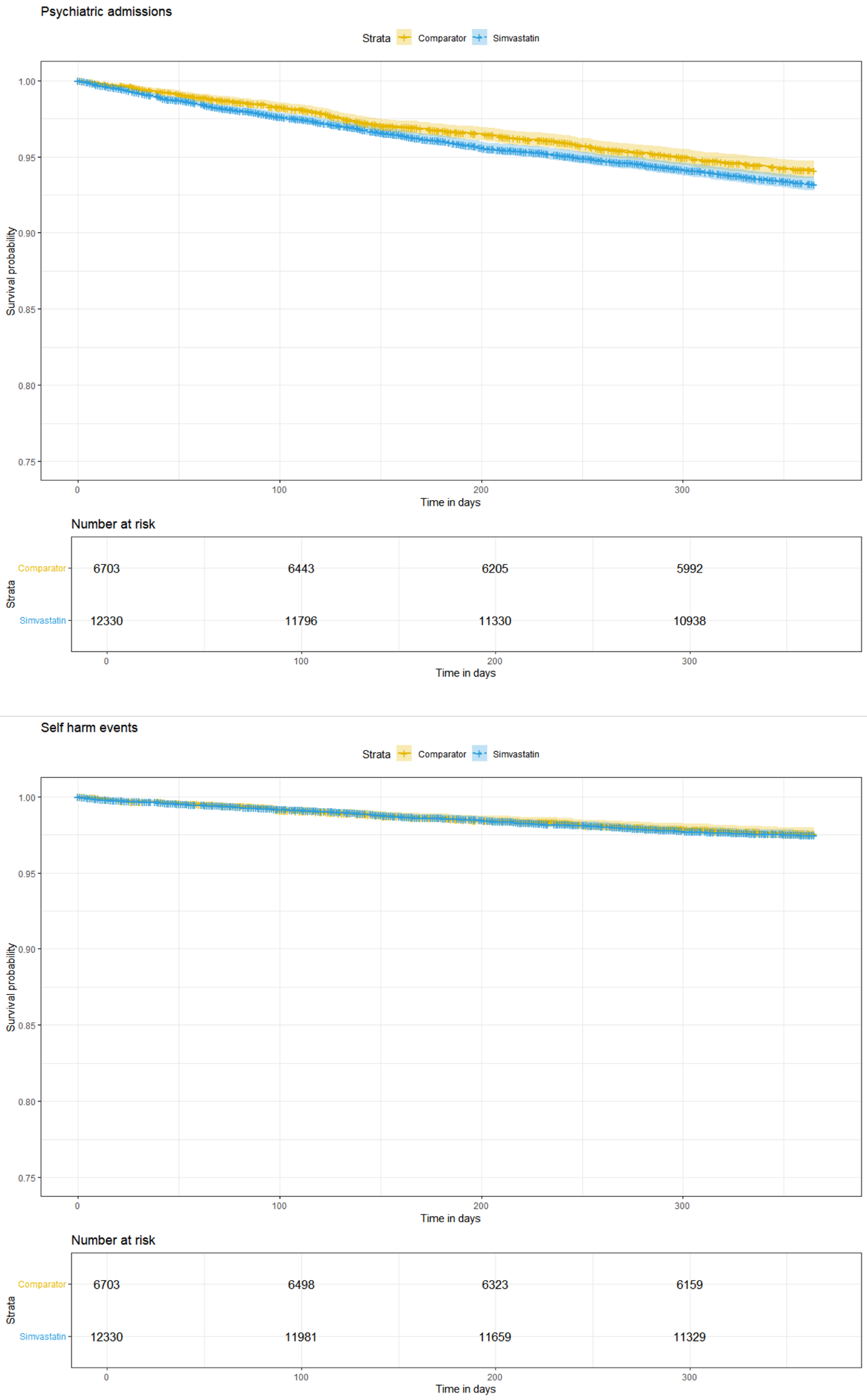

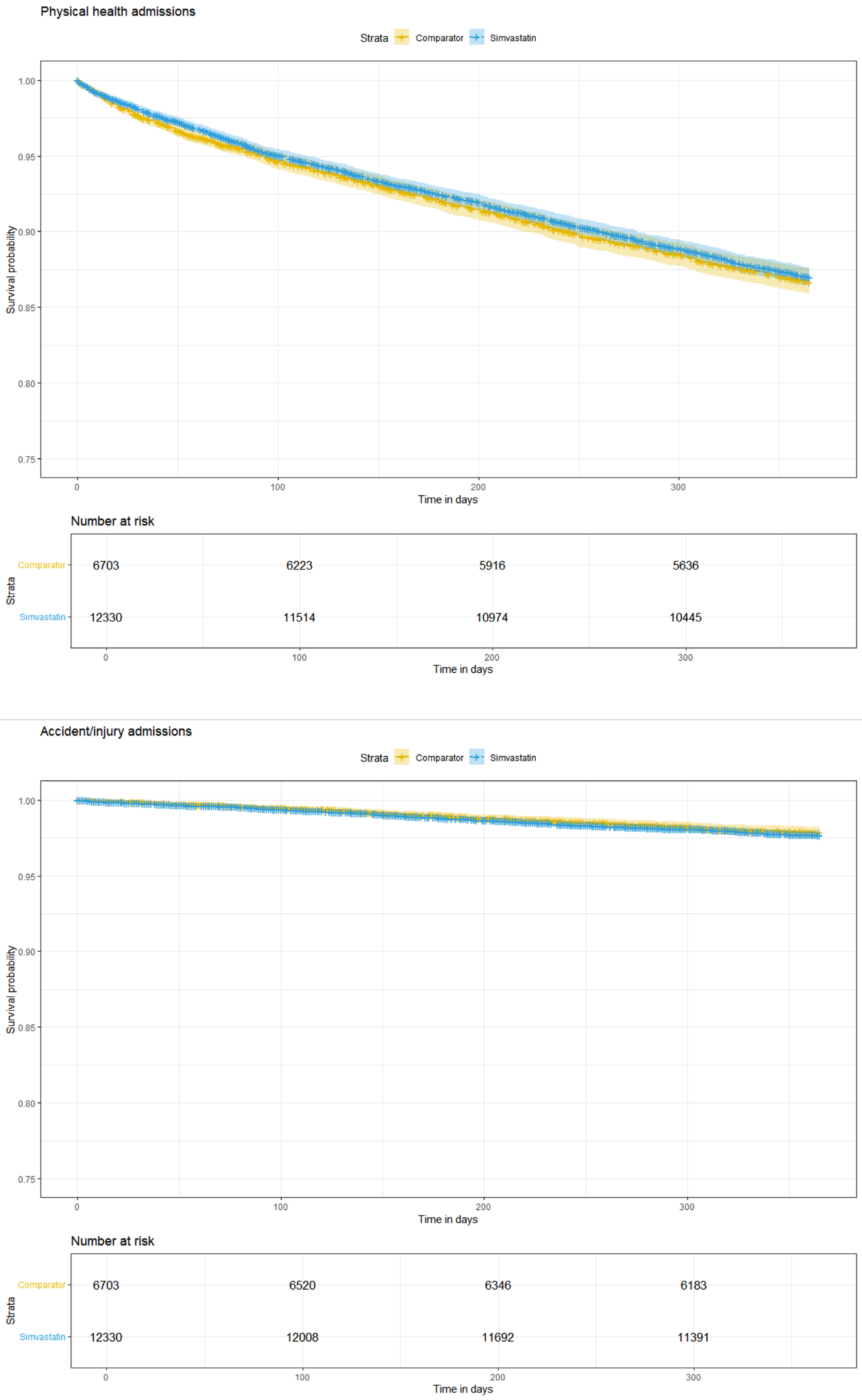

##### Supplementary Figure 3: Schoenfeld residual and Kaplan Meier plots, Trial 2A

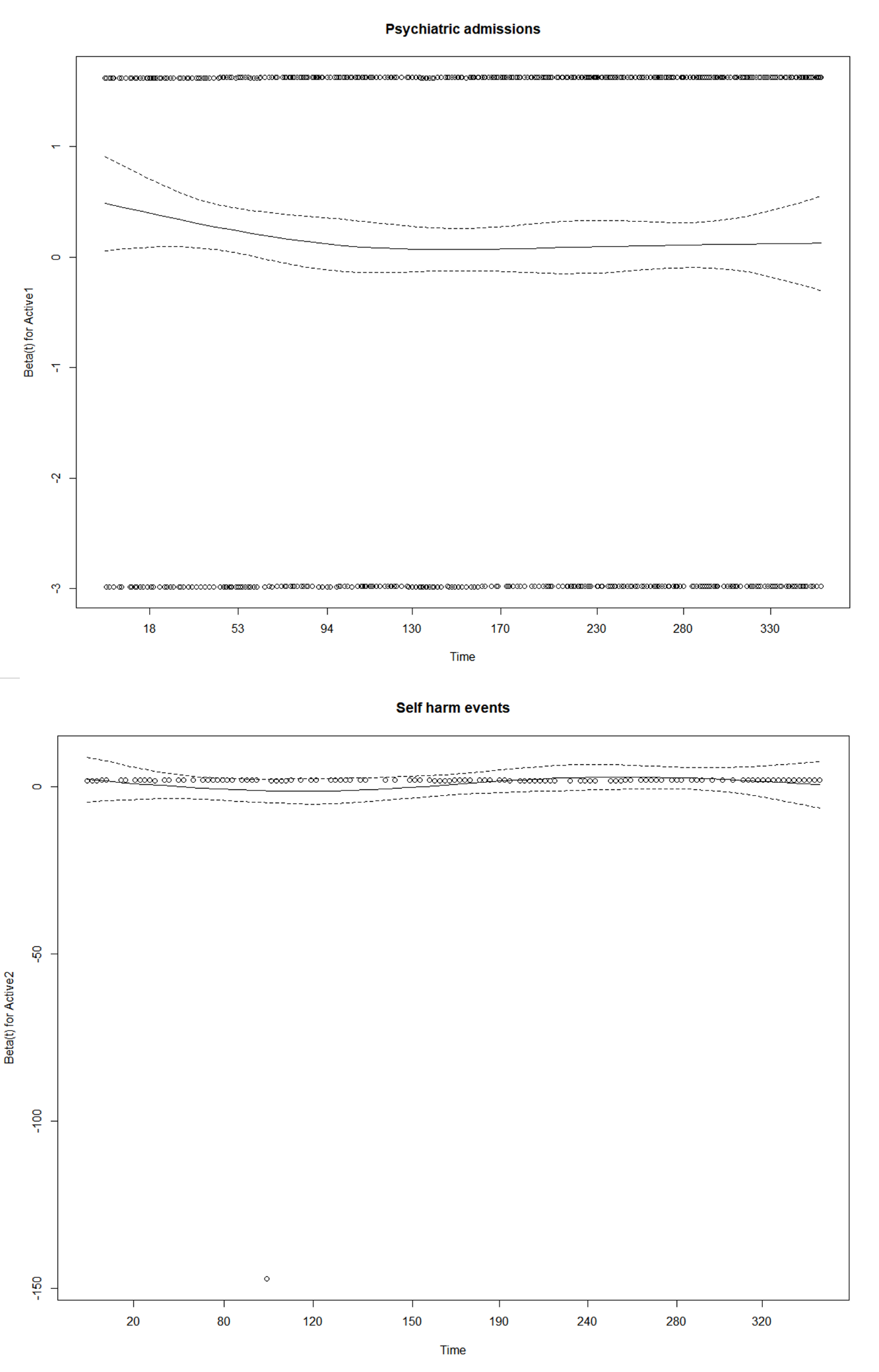

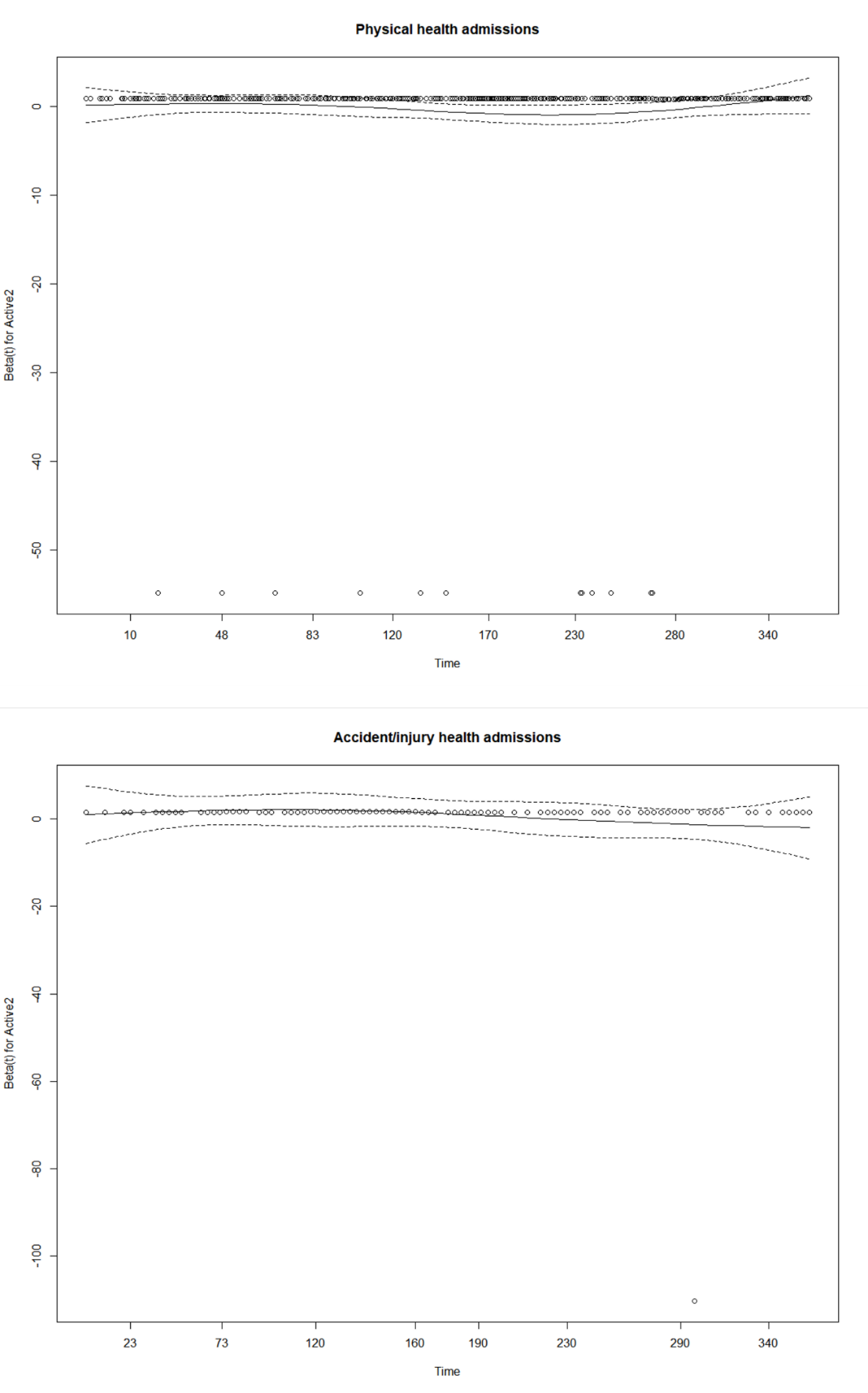

Psychiatric admissions: p=0.72; self-harm: p=0.39; physical health admissions: p=0.72, accident

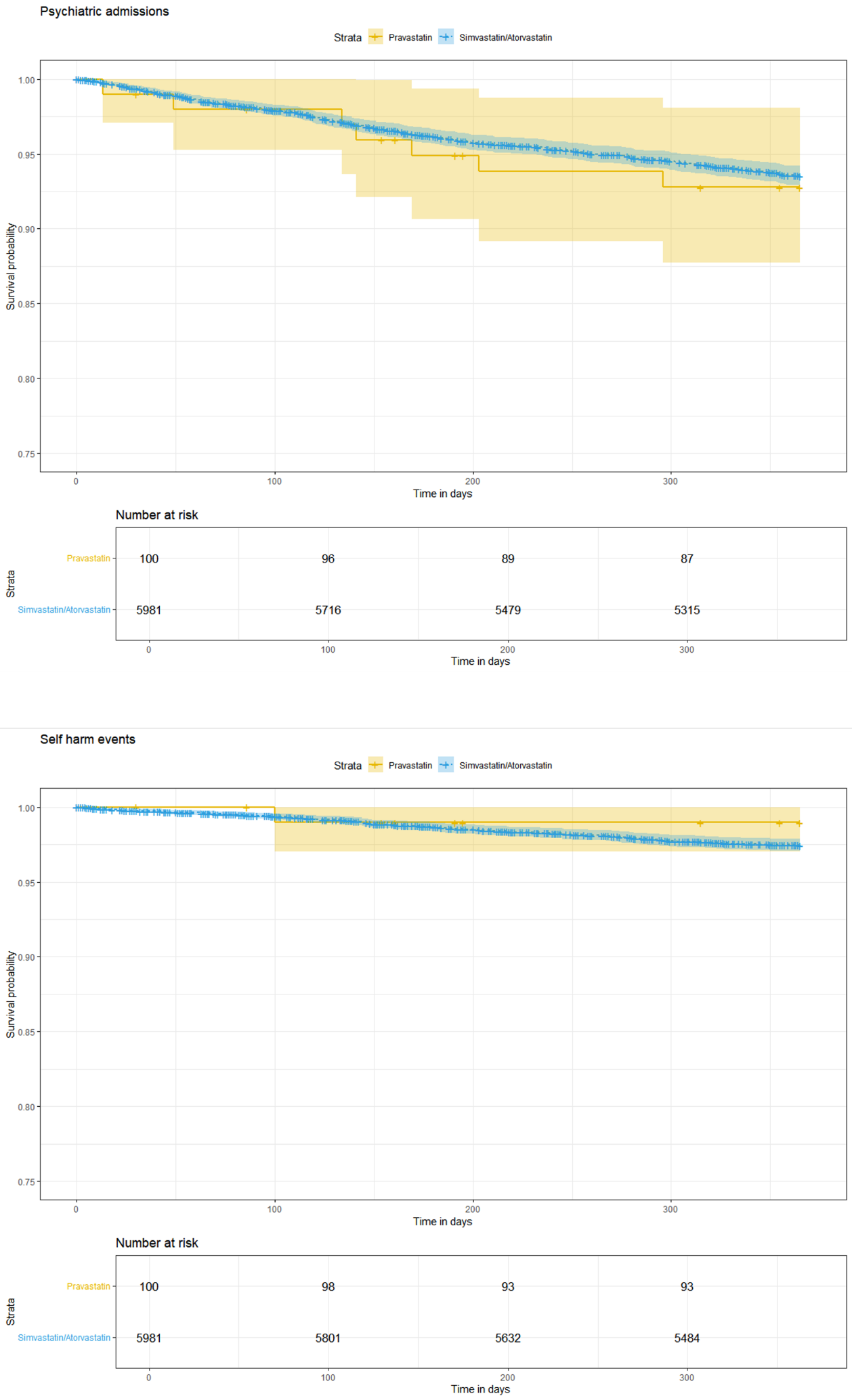

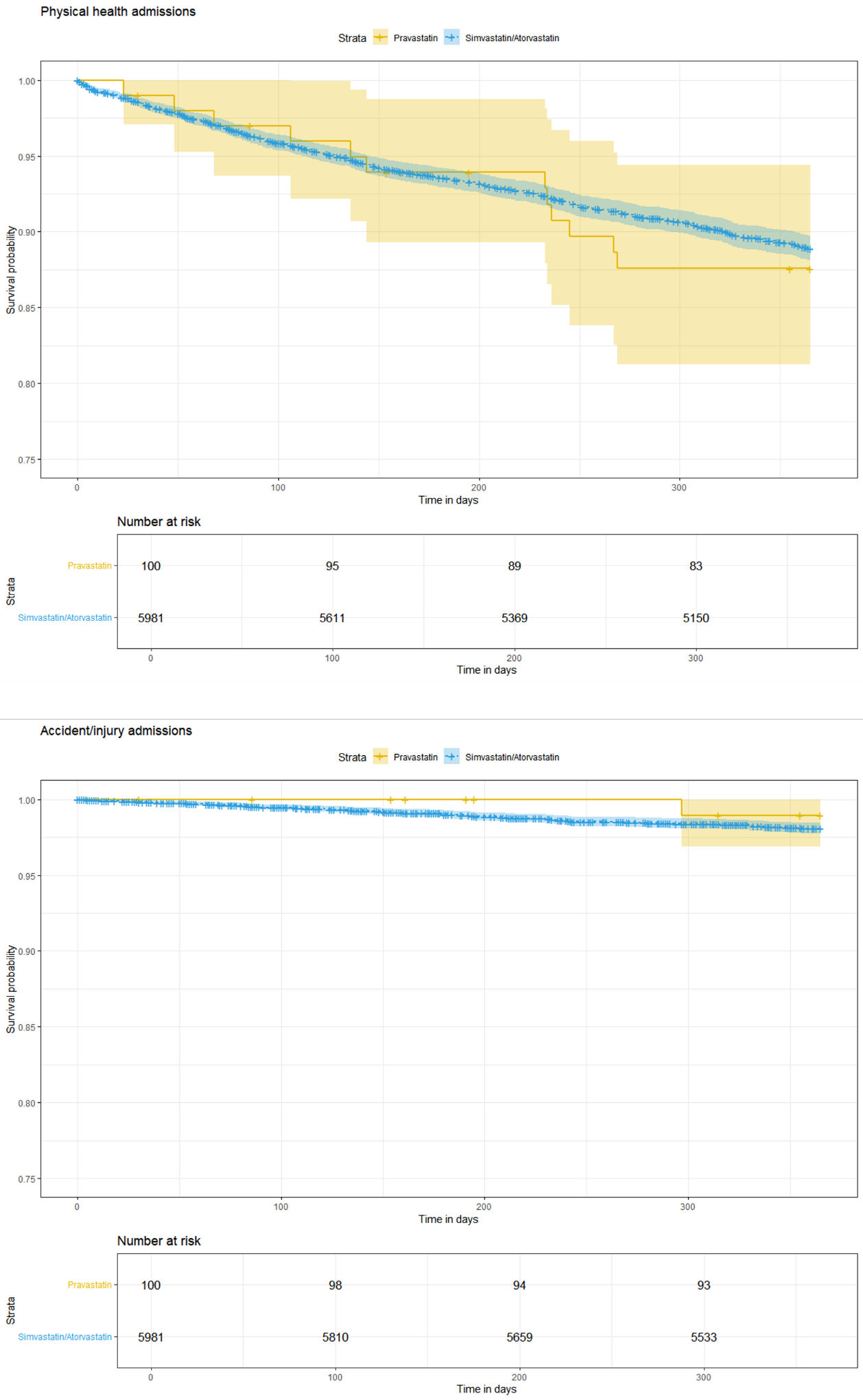

##### Supplementary Figure 4: Schoenfeld residual and Kaplan Meier plots, Trial 2B

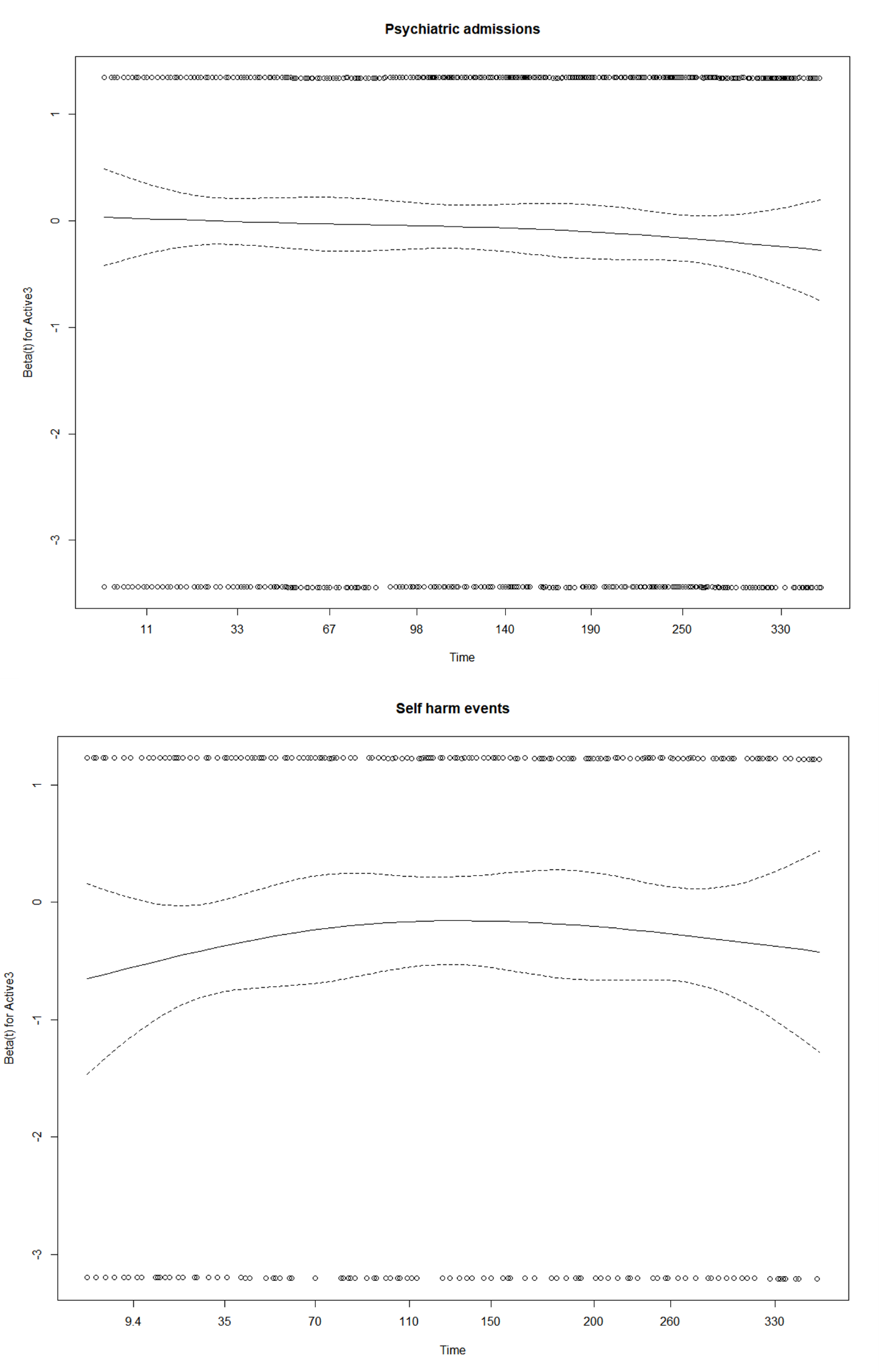

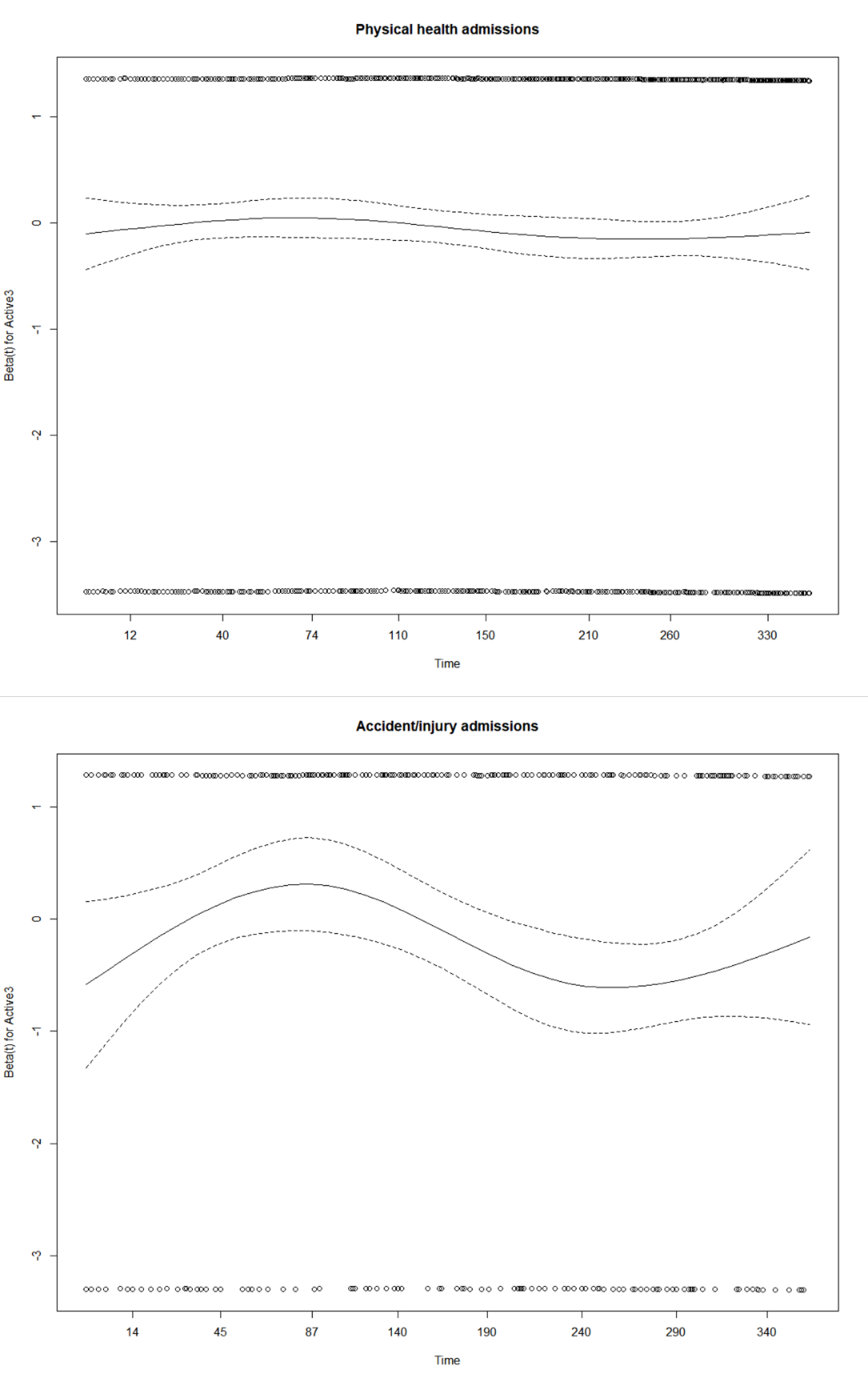

Psychiatric admissions: p=0.24; self-harm: p=0.64; physical health admissions: p=0.31; accident and injury admissions: p=0.13

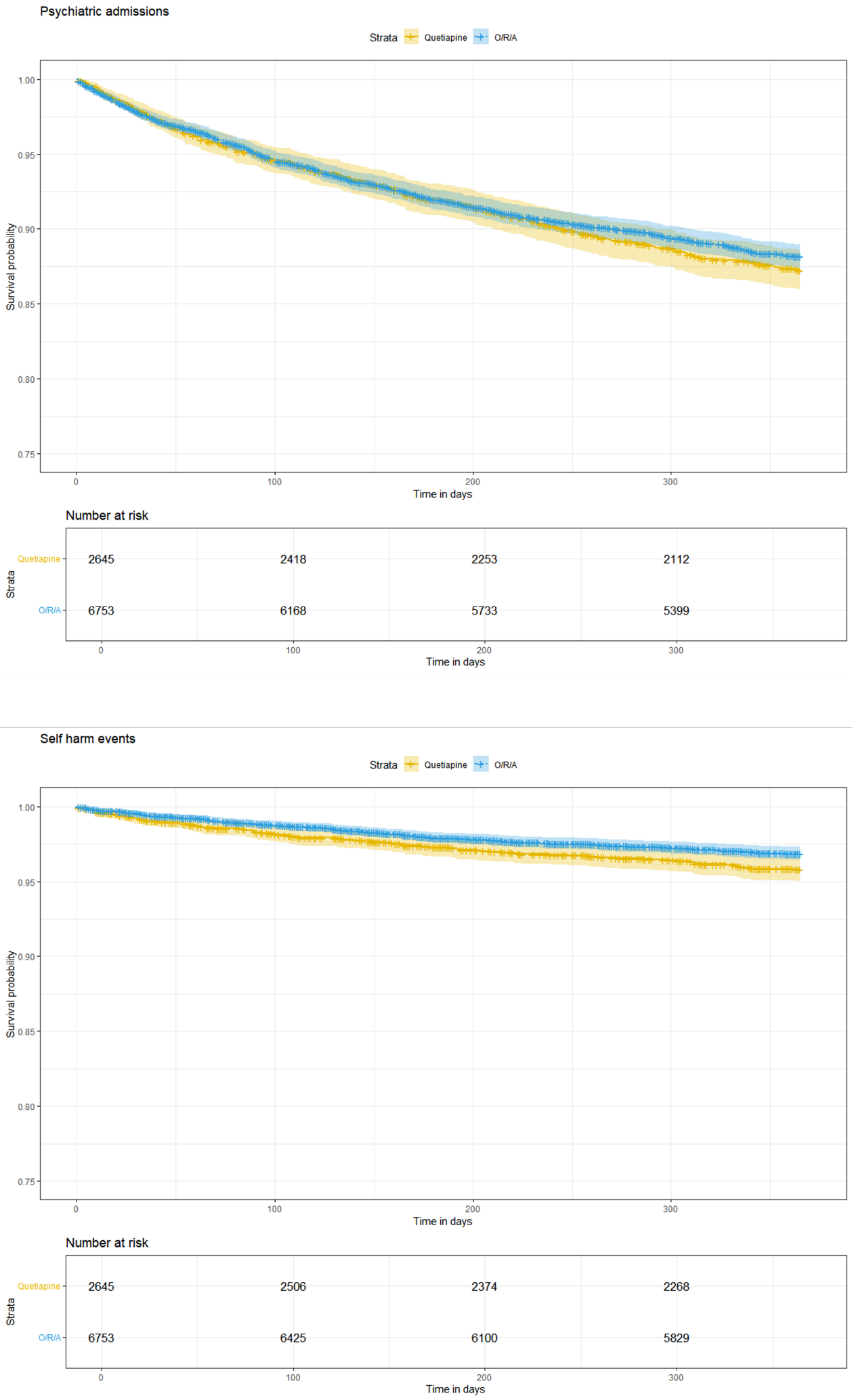

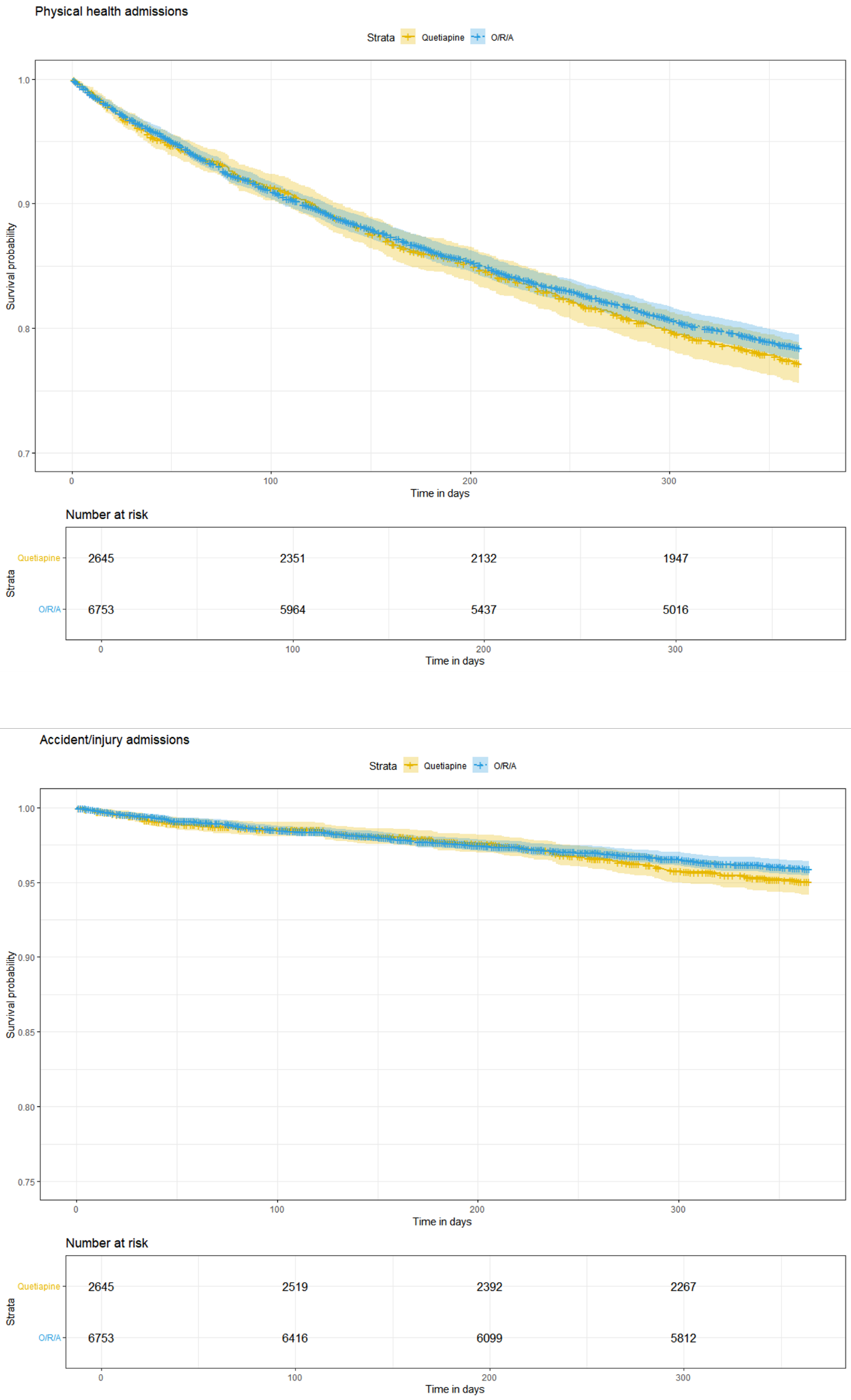

##### Supplementary methods

**Models of count data**

We tested Poisson models of each outcome at 12 months for evidence of zero inflation and over dispersion. For all trials, the data was over dispersed but not zero-inflated for all outcomes other than self-harm in trial 1 and physical health admissions in trial 2B These two outcomes were zero-inflated as well as over dispersed. We therefore used a negative binomial model to account for over dispersion. There was no evidence of zero-inflation when using the negative binomial models.

**Inverse probability weighting**

We performed inverse probability weighting with stabilised weights without trimming. We used a threshold for potential imbalance of >0.1^a^.

For Trial 1, the only variable to fall outside the 0.1 threshold was statin dose, with high dose statins more commonly prescribed in the comparator arm and moderate dose in the active arm. We ran IPW models with and without additional adjustment for this variable (supplementary methods figure 1).

For Trial 2A, the number of patients in the comparator arm was small. Furthermore, these patients were markedly different to the active arm. We failed to balance covariates for this trial. This led to an effective sample size of 3.53 patients in the active arm (supplementary methods figure 2).

For Trial 2B, all variables were within the 0.1 threshold following weighting (supplementary methods figure 3).

**Per protocol analysis**

For trial 1, follow up time ended at the earliest of: end of follow up as per the main analysis, discontinuation of antipsychotics/mood stabilisers or statins (defined as a gap of at least 90 days), or for the active arm, switching to a statin other than simvastatin, and the control arm switching to a statin other than atorvastatin, pravastatin or rosuvastatin.

For trial 2A, follow up time ended at the earliest of: end of follow up as per the main analysis, discontinuation of aripiprazole, risperidone or olanzapine (defined as a gap of at least 90 days); discontinuation of statins (defined as a gap of at least 90 days), or for the active arm, prescription of a statin other than simvastatin, and the control arm prescription of a statin other than atorvastatin, pravastatin or rosuvastatin. We did not terminate follow up at the prescription of antipsychotics not included in the study, provided aripiprazole, risperidone or olanzapine were continued.

For trial 2B, follow up time ended at the earliest of: end of follow up as per the main analysis, discontinuation of simvastatin or atorvastatin (defined as a gap of at least 90 days); discontinuation of risperidone, olanzapine, aripiprazole or quetiapine (defined as a gap of at least 90 days), or for the active arm, any prescription of quetiapine, and the control arm switching to risperidone, olanzapine or aripiprazole. We did not terminate follow up at the prescription of antipsychotics not included in the study, provided a patient did not initiate an antipsychotic in the other study arm.

a: Austin, P.C. and E.A. Stuart. Moving towards best practice when using inverse probability of treatment weighting (IPTW) using the propensity score to estimate causal treatment effects in observational studies. Stat Med, 2015. 34(28): p. 3661-79.

Supplementary methods Figure 1: Covariate balance before and after imputation for Trial 1

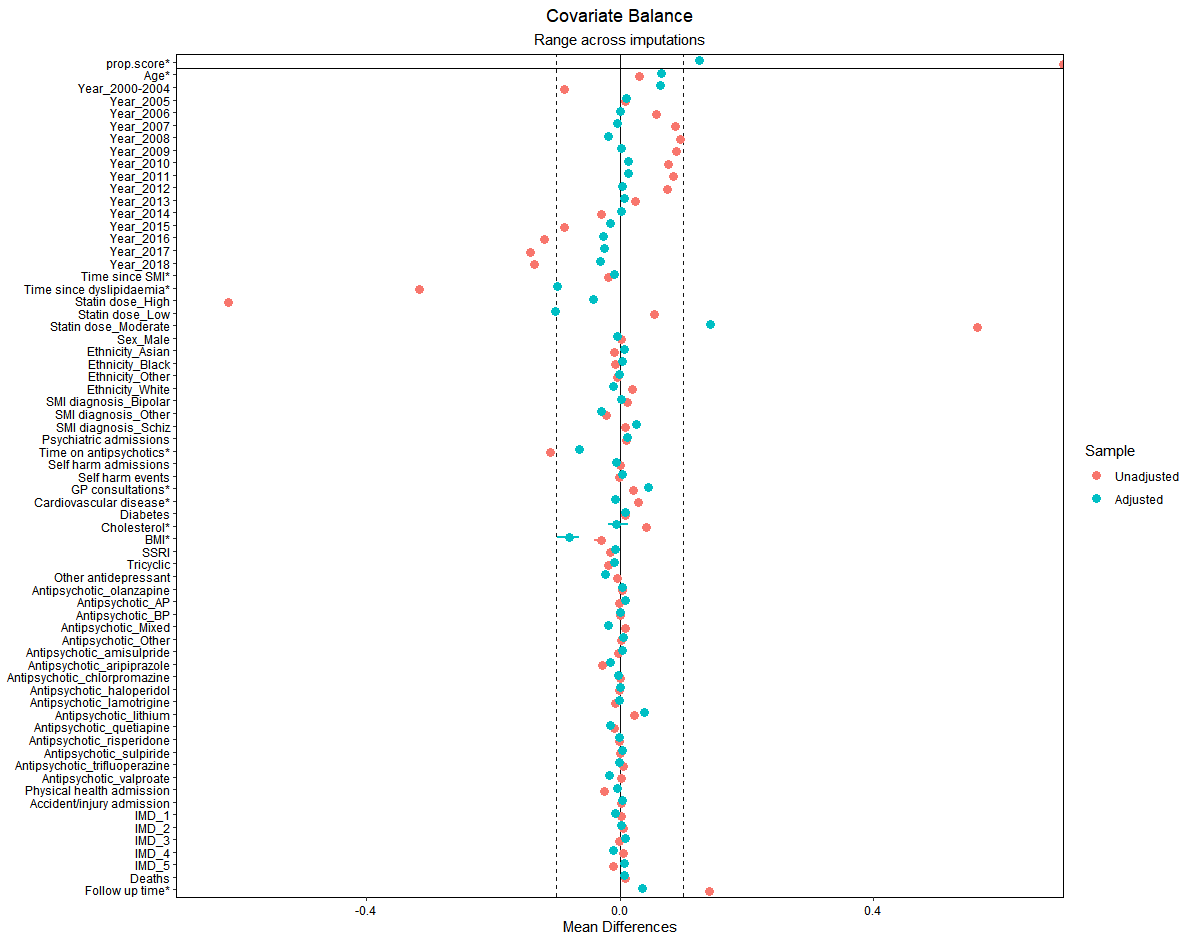

Supplementary methods Figure 2: Covariate balance before and after imputation for Trial 2A

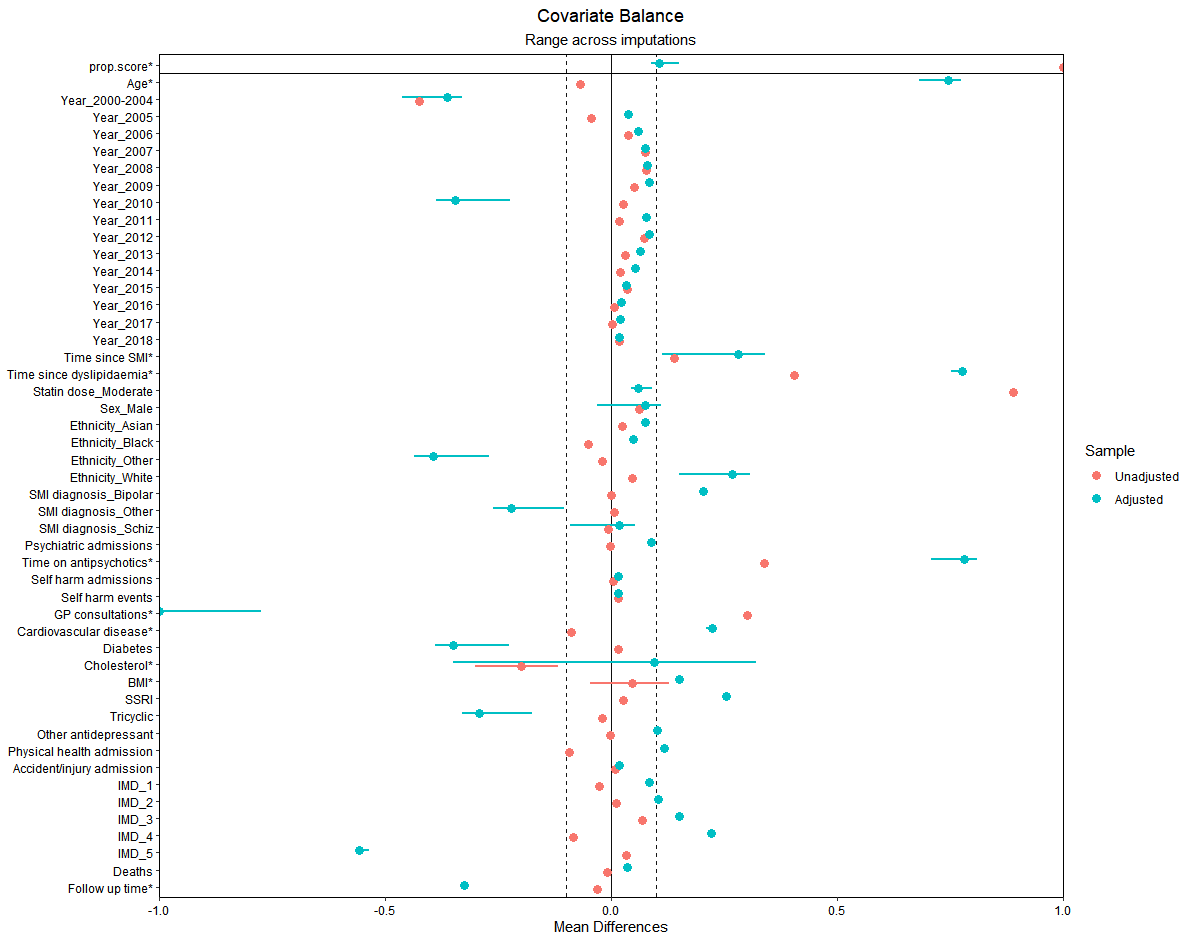

Supplementary methods Figure 3: Covariate balance before and after imputation for Trial 2B

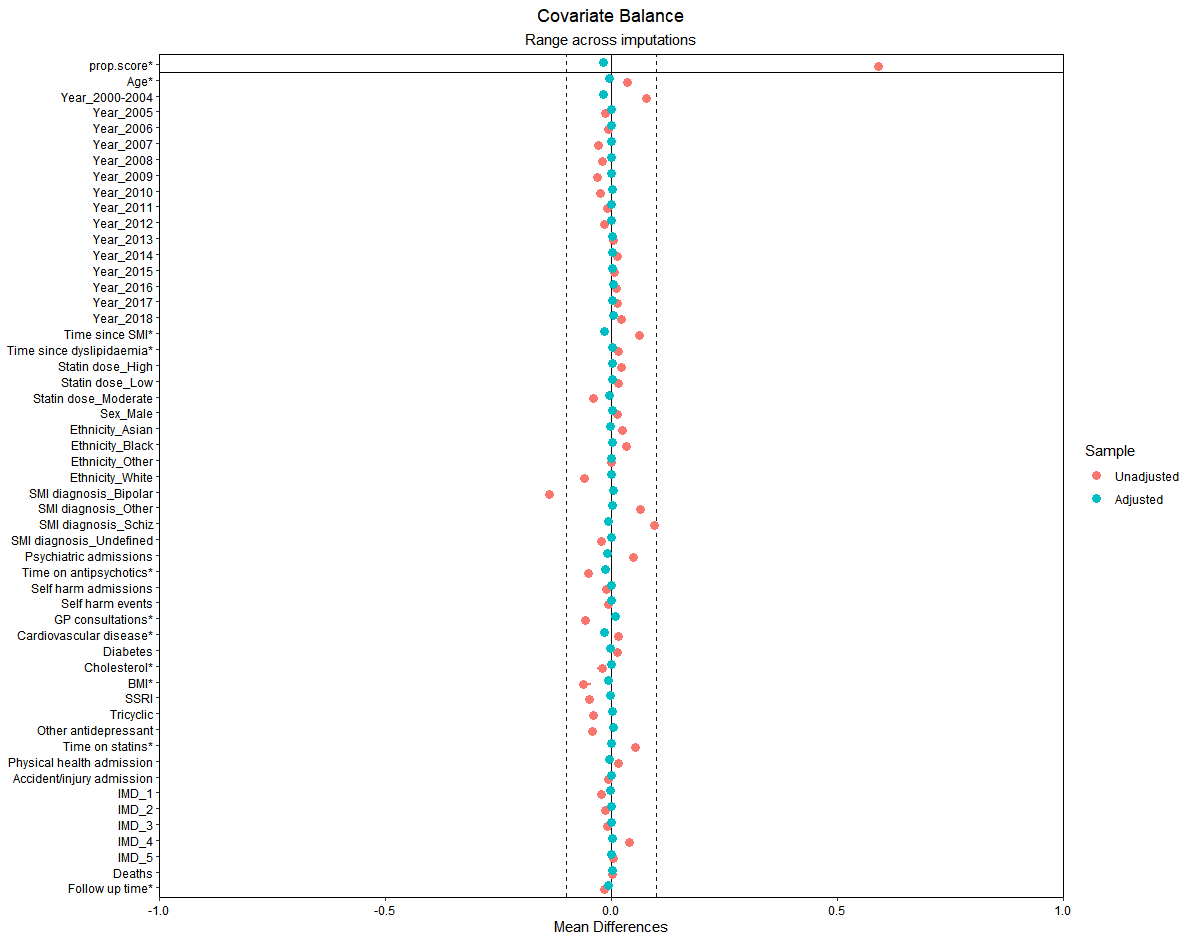
